# SLC39A8.p.(Ala391Thr) is associated with poorer cognitive ability: a cross-sectional study of schizophrenia and the general UK population

**DOI:** 10.1101/2024.09.18.24313865

**Authors:** Sophie E Smart, Sophie E. Legge, Eilidh Fenner, Antonio F. Pardiñas, Grace Woolway, Amy J. Lynham, Valentina Escott-Price, Jeremy Hall, Lawrence Wilkinson, Peter Holmans, Michael C. O’Donovan, Michael J. Owen, James T.R. Walters

## Abstract

The missense SNP NC_000004.12:g.102267552C>T (SLC39A8.p.(Ala391Thr), rs13107325) in *SLC39A8,* which encodes a zinc transporter, has been linked to schizophrenia and is the likely causal variant for one of the genome-wide association loci associated with the disorder.

We tested whether the schizophrenia-risk allele at p.(Ala391Thr) was associated with schizophrenia-related phenotypes, including positive, negative, and disorganised symptoms, cognitive ability, educational attainment, and age of psychosis onset, within three schizophrenia cohorts (combined N=1,232) and, with equivalent phenotypes, in a sample of population controls (UK Biobank, N=355,069). We used regression analyses controlling for age, sex, and population stratification.

Within the schizophrenia cohorts, after correction for multiple testing, p.(Ala391Thr) was not significantly associated with any schizophrenia-related phenotypes. In the unaffected participants from the UK Biobank, the schizophrenia-risk allele at p.(Ala391Thr) was associated with significantly poorer cognitive ability and fluid intelligence, a lower probability of obtaining GCSEs or a degree-level qualification, and fewer years in education. There was no association between p.(Ala391Thr) and self-reported psychotic experiences in this cohort.

The schizophrenia-risk allele was associated with poorer cognitive ability, but not psychotic experiences, in a volunteer sample drawn from of the general population. To determine whether p.(Ala391Thr) is associated with cognitive phenotypes in people with schizophrenia, and to understand the role of p.(Ala391Thr) in the aetiology of cognitive impairment in schizophrenia, larger independent samples are required.

## Introduction

The gene *SLC39A8* encodes metal cation symporter ZIP8, which enables the movement of at least five trace elements (manganese, zinc, iron, selenium, and cobalt) from outside the cell into the cytosol. *SLC39* genes are highly conserved and are critical for fundamental life processes throughout the lifespan. *SLC39A8* contains a missense single nucleotide polymorphism (SNP), NC_000004.12:g.102267552C>T (SLC39A8.p.(Ala391Thr), rs13107325), in which alternative alleles encode alanine (Ala; C allele) or threonine (Thr; T allele) on chromosome 4 (1). In vitro, the minor allele (T) leads to reduced uptake of manganese and cadmium (2), the latter a toxic environmental pollutant, and decreased synaptic uptake of zinc (3).

p.(Ala391Thr) is associated with an increased risk of schizophrenia, a heterogeneous disorder whose presentation includes delusions, hallucinations, behavioural disturbance, social withdrawal, and cognitive impairment. In the most recent genome-wide association study (GWAS), of 69,369 schizophrenia cases and 236,642 controls, the minor allele was associated with increased the risk of schizophrenia (odds ratio=1.17, P=2.9×10^-21^) (4). Furthermore, fine-mapping suggested that p.(Ala391Thr) was likely to be the causal variant underpinning the association with the genomic region containing *SLC39A8*, the SNP having a posterior probability of being causally associated with schizophrenia of 99.20%. Only nine out of 255 loci were fine-mapped to one causal variant (4), only two of these were missense variants, and only p.(Ala391Thr) had a Combined Annotation-Dependent Depletion (CADD) score of 22 making it one of the 1% most deleterious variants in the genome.

In non-psychiatric populations, p.(Ala391Thr) has not been associated with reported psychotic experiences (Z = 0.99, P = 0.889) (5), but the schizophrenia risk allele has been associated with lower intelligence (Z = -9.49, P = 2.23E-21) (6), fewer years in education (Beta = -0.02, P = 1.08E-13) (7), and, using a GWAS-by-subtraction design, with the cognitive aspect of educational attainment (NonCog, Z = 2.78, P = 0.005; Cog, Z = -9.87, P = 5.59E-23) (8). To date, no studies have examined the relationship between p.(Ala391Thr) and the clinical presentation or cognitive ability of people with schizophrenia.

Our primary aim was to test whether schizophrenia-risk alleles at p.(Ala391Thr) are associated with phenotypes that capture the clinical presentation of schizophrenia. We hypothesised that the schizophrenia-risk allele (T) at p.(Ala391Thr) would be associated with a more severe clinical presentation, poorer cognitive ability and educational attainment, and younger age of onset. We expanded the analysis to test for similar phenotypic associations in a large sample of the general population (UK Biobank unaffected controls). Given that *SLC39A8* is also constrained for loss of function and missense variants (Genome Aggregation Database [gnomAD] v2.1.1; 9), we used the UK Biobank to tested for phenotypic associations with rare protein-truncating and deleterious missense variants within *SLC39A8*.

## Methods

This study followed the guidelines outlined in the STrengthening the REporting of Genetic Association Studies (STREGA) (10), an extension of the STROBE Statement (checklist in the Supplementary Material).

We analysed data from three cohorts recruited at Cardiff University and data from the UK Biobank. Full details on how samples were ascertained and how phenotypes were measured, calculated, and standardised, can be found in the Supplementary Material. A brief overview is provided below.

### Cardiff Schizophrenia Samples

#### Participants

Participants were recruited into three cross-sectional studies, all of which have been previously described: CardiffCOGS (11), Cardiff F-series (12) and Cardiff Affected-Sibs (13) samples. The Cardiff Affected-Sibs sample includes a single affected individual from families with two or more siblings diagnosed with schizophrenia or schizoaffective disorder. All participants were recruited from in-patient, voluntary sector, and community mental health services across the UK and underwent a clinical research interview based on the Schedules for Clinical Assessment in Neuropsychiatry (SCAN), with further information available from clinical records. For these analyses, we only retained participants who met DSM-IV or ICD-10 criteria for a diagnosis of schizophrenia or schizoaffective disorder, depressed type.

#### Phenotypes

The Scale for the Assessment of Positive Symptoms (SAPS) and the Scale for the Assessment of Negative Symptoms (SANS) were scored on a lifetime worst basis using information from the SCAN interview and lifetime psychiatric clinical records. These records were also used to ascertain age at psychosis onset. The Measurement and Treatment Research to Improve Cognition in Schizophrenia (MATRICS) Consensus Cognitive Battery was used as a measure of current cognitive ability in the CardiffCOGS sample. As described in Legge, Cardno (14), a confirmatory factor analysis (CFA) framework was used to estimate phenotype-derived factor scores from the symptom and cognitive ratings, which we refer to as symptom dimensions. Using all three samples, the best model had three symptom dimensions: positive symptoms (SAPS global hallucinations and SAPS global delusions), negative symptoms of diminished expressivity (SANS global affective flattening and SANS global alogia), and disorganised symptoms (SAPS global positive formal thought disorder and SANS inappropriate affect). As CardiffCOGS was the only sample to have data on cognitive ability, a second CFA framework, using only this sample, was used to estimate phenotype-derived factor scores from the MATRICS domain scores as well as the symptom ratings. The best model had five dimensions: positive symptoms (as above), negative symptoms of diminished expressivity (as above), disorganised symptoms (as above), negative symptoms of motivation and pleasure (SANS global anhedonia/asociality and SANS global avolition/apathy), and cognitive ability (all MATRICS domains apart from social cognition).

As well as the cognitive domain, which captures current cognitive ability, premorbid IQ was assessed in the CardiffCOGS sample using the National Adult Reading Test (NART). Educational attainment was measured across all three samples using years in education and highest educational qualification (General Certificate of Secondary Education [GCSE]/no GCSE, note GCSEs are taken by most UK pupils upon the completion of compulsory education [from 1972-2013 this was at age 16], and degree/no degree).

#### Genotypes

Genome-wide SNP data for the three samples were curated and harmonised as part of DRAGON-Data (15) (see Supplementary Material). Genotypes for p.(Ala391Thr) (INFO=0.996) were extracted for each participant. Principal components were calculated using pruned SNPs.

Genetic ancestry probabilities were calculated using Ancestry Informative Markers (AIMs) derived from the Allen Ancient DNA Resource reference panel, linear discriminant analysis (LDA), and biogeographical categories defined by Huddart, Fohner (16). AIMs are genetic variants with highly divergent allele frequencies across biogeographical genetic ancestries, which can capture genetic associations between an individual and a particular (sub)continental population (17). Ancestry groups were determined by assigning individuals to their most probable biogeographical category as inferred by LDA (determined using Youden’s index as optimality criterion), with individuals not meeting a probability threshold for any category being assigned to an “admixed” group. The schizophrenia risk allele is thought to have been under recent positive selection in Europeans (18); in Phase 3 of the 1000 Genomes Project, the schizophrenia allele is present in around 8% of European samples but is reported to be almost absent from other populations (1). In our data, only Europeans carried two copies of the schizophrenia risk allele. However, to increase our sample size and improve the generalisability of our findings, we restricted our sample to participants from ancestries where the schizophrenia-risk allele was present in at least one individual: African American/Afro-Caribbean (0.48%), Central/South Asian (1.06%), European (98.21%), and Middle Eastern/North African (0.24%) (note the Middle Eastern/North African ancestry is referred to as Near Eastern in Huddart, Fohner (16)).

### UK Biobank

#### Participants

Participants were from the UK Biobank (UKBB), a large-scale biomedical database of individuals aged between 40-69 who were recruited from across the UK (19). Participants underwent extensive phenotyping. All UKBB field IDs used in this study are reported in the Supplementary Material. For these analyses, we removed participants with a psychotic spectrum disorder (ICD-10 F20-F29, see Supplementary Material).

#### Phenotypes

An abridged version of the Composite International Diagnostic Interview psychosis module (lifetime version) was used in the online follow-up mental health questionnaire. As in Legge, Jones (5), we derived three overlapping binary variables: (i) any psychotic experience defined as a positive response to any of the four symptom questions; (ii) a distressing psychotic experience, defined as any psychotic experience that was rated as “a bit,” “quite,” or “very” distressing; and (iii) multiple occurrences of psychotic experiences, defined as any psychotic experience that occurred on more than one occasion. The comparator group for these three variables was comprised of individuals who provided a negative response to all four psychotic experience symptom questions (see Supplementary Material). We also looked at delusions of persecution alone (UKBB Field 20468: “Ever believed in an un-real conspiracy against self”) as previous work has suggested that this phenotype in the UKBB is particularly enriched for genetic liability for schizophrenia (5).

Cognitive ability was measured using a general intelligence factor, *g*, which is considered a reliable measure of cognitive ability. As in Fawns-Ritchie and Deary (20), *g* was calculated using principal component analysis (PCA). Four cognitive tests went into the PCA (numeric memory, reaction time, pairs matching, and trail making test (TMT) B, see Supplementary Material) and the first PC was considered an estimate of g. A positive *g* score represents better cognitive performance. Alongside the *g* score, the cognitive test of verbal and numerical reasoning, also referred to as the test for fluid intelligence, was used to measure current cognitive ability. Educational attainment was measured using years in education (for those without a college or university degree) and highest educational qualification (GCSEs/no GCSEs and degree/no degree).

#### Genotypes

Genotype data were curated by the UKBB (21) and as described in Leonenko, Baker (22) (see Supplementary Material). The genotypes for p.(Ala391Thr) (INFO=1.00) were extracted for each participant. Principal components provided by the UKBB were used. We restricted our sample to participants who self-reported White British or Irish ancestry then used the first five principal components to identify a subsample of participants that was relatively genetically homogenous. As in Legge, Jones (5), we computed a Minimum Covariance Determinant (MCD) estimator of location and scatter for each participant, and used these to define a hyper-ellipsoid in a multi-dimensional space that contains the majority of MCD points. We used this hyper-ellipsoid to include participants within the 90^th^ percentile of the MCD distance (see Figure 9 in the Supplementary Material).

#### Rare Variants

Full methods of rare variant calls are reported in Fenner, Holmans (23). In brief, exome sequencing data released for 200,619 UKBB participants in October 2020 were used for the current study. Variants were annotated in Hail using Ensembl’s VEP. Protein-truncating variants (PTVs) were defined as splice acceptor, splice donor, stop-gain or frameshift variants that were annotated as high confidence for causing loss of protein function by Loss-Of-Function Transcript Effect Estimator (LoFTEE; 9). Deleterious missense variants were defined as missense variants with a Rare Exome Variant Ensemble Learner score (REVEL; 24) > 0.75. PTVs and missense variants were grouped together into one ‘damaging’ category for analysis. Rare variants were defined as those with allele counts ≤ 5 in the sample reported in Fenner, Holmans (23). We included, as a covariate, the burden of synonymous variants that had allele counts ≤ 5 in the sample reported in Fenner, Holmans (23).

### Statistical Analysis

Each phenotype was regressed onto p.(Ala391Thr) using linear or logistic regression models. Sex, age at interview, the first five genetic principal components (PCs), and any further PCs which were associated with p.(Ala391Thr) (PC 6 and 9 in the Cardiff F-Series sample) were included as covariates. For the UKBB data, the first ten genetic PCs and genotype batch were included as covariates. For models where a measure of cognitive ability in the UKBB was used as the outcome, age at interview squared was also included as a covariate (25), this variable was centred before being transformed to prevent the occurrence of collinearity (26). For the Cardiff schizophrenia samples, missing phenotype data were imputed, for each sample separately, using multiple imputation by chained equations (MICE; 100 imputations, 10 iterations in the burn-in period). All regression analyses were run in each of the 100 imputed datasets separately and then pooled using Rubin’s rules (27). Model assumptions were checked using the R package ‘performance’ and discussed in the Supplementary Material: for models were there was evidence of multicollinearity (variance inflation factor [VIF] ≥ 10), we removed PCs from the model until the VIF for p.(Ala391Thr) was < 10, and for models where there was evidence of heteroscedasticity (Breusch-Pagan Test P-value < 0.01), we recalculated standard errors, confidence intervals, and P-values using Eicker-Huber-White robust “HC2” standard errors. For the Cardiff schizophrenia samples, beta values or log(odds) were meta-analysed, and weighted using their standard errors, using the R package ‘metafor’ using a fixed-effect inverse-variance weighted model. We corrected for multiple testing of phenotypes in the meta-analysis, and within each sample, using the Benjamini–Hochberg False Discovery Rate (FDR) method and used an alpha level of ≤ .05 for the adjusted p-values to determine statistical significance. For the rare variant analysis, each of the phenotypes was regressed against the number of rare variants, and adjusted for the number of rare synonymous variants each person carries in *SLC39A8*, age at interview (age at interview squared for *g* and FI), sex, exome data PC1-10, and sequencing batch.

## Results

Phenotype means and proportions, stratified by p.(Ala391Thr) schizophrenia-risk allele count, are presented for each sample in the Supplementary Material.

### Cardiff Schizophrenia Samples

Data were available for 662 participants from CardiffCOGS (mean age at interview [AAI]=43.31 years, standard deviation [sd]=12.11; 65.75% male), 422 from Cardiff F-Series (mean AAI=42.11 years, sd=14.08; 70.66% male) and 148 from Cardiff SibPairs (mean AAI=41.68 years, sd=12.93; 66.21% male).

p.(Ala391Thr) was not associated with positive symptoms, negative symptoms, or disorganised symptoms derived in either CFA model (Table 1 and Table 2). p.(Ala391Thr) was not associated with current cognitive ability in CardiffCOGS, however, it was nominally associated with a lower NART IQ score but this association did not survive correction for multiple testing (Table 2). Although I^2^ should be interpreted cautiously in meta-analyses of only a few studies (28), there does not appear to be evidence of between-study heterogeneity after meta-analysing phenotypes which were present in all three samples.

### UK Biobank

Data were available for N=355,069 participants from UKBB who did not have a psychotic spectrum disorder (mean AAI=56.92 years, sd=7.96; 46.25% male), of which N=354,509 indicated that they were willing to attempt the cognitive tests and N=116,935 completed the online follow-up mental health questionnaire.

p.(Ala391Thr) was not associated with reported psychotic experiences. However, the schizophrenia-risk allele was associated with a lower *g* score (β = -0.05; 95% CI, -0.07 to - 0.02; FDR-adjusted p-value=8.61 x 10^-5^), a lower fluid intelligence score (β=-0.05; 95% CI, - 0.07 to -0.04; FDR-adjusted p-value=3.35 x 10^-10^), a lower likelihood of obtaining GCSEs (OR=0.96; 95% CI, 0.94-0.98; FDR-adjusted p-value=6.21 x 10^-5^) or a degree-level qualification (OR=0.95; 95% CI, 0.93-0.97; FDR-adjusted p-value=6.03 x10^-6^) and fewer years in education (β=-0.03; 95% CI, -0.05 to 0.00; FDR-adjusted p-value=0.035) (Table 3).

Rare variant calls were available for N=134,370 (37.84%) of the participants included in the current study. N=46 carried rare synonymous variants and N=13 carried rare damaging variants (N=6 PTVs and N=7 missense carriers; mean AAI=55.54 years, sd=9.92; 46.15% male). Most rare variant carriers had missing phenotype data. Rare variants in *SLC39A8* were nominally associated with lower *g* (N rare variants=2; β = -1.67; 95% CI, -2.93 to -0.42; P-value=0.009) and lower fluid intelligence (N rare variants=1; β = -2.31; 95% CI, -4.25 to - 0.38; P-value=0.019) but this association did not survive correction for multiple testing (Table 4). None of the rare variant carriers had a diagnosis of Intellectual Disability (ICD-10 Codes F70, F71, F72, F78, or F79 identified from the hospital, death, and primary care records).

## Discussion

In this study, we found that the schizophrenia-risk allele at p.(Ala391Thr) was associated with poorer cognitive ability, but not psychotic experiences, in a volunteer sample, drawn from the general population, without psychotic spectrum disorders. The schizophrenia-risk allele was also nominally associated with lower premorbid IQ in patients with schizophrenia; larger independent samples are required to confirm this result. Exploratory analysis of rare variants in *SLC39A8*, in our subsample of the UKBB, suggested that rare variants are nominally associated with poorer cognition but was not decisive due to the low number of rare variant carriers with phenotype data.

Although understanding the potential pathophysiological mechanisms of variants identified by GWAS is challenging, there is an increasing focus on p.(Ala391Thr) in the context of the aetiology of schizophrenia (1) and some proposed mechanisms are also thought to play a role in cognition. p.(Ala391Thr) is thought to lead to synaptic glutamate receptor hypofunction, in part, because of downregulated surface localisation of GluA1, GluA2/3, GluN1, and GluN2A (3). The latter are subunits of the *N*-methyl-d-aspartate (NMDA) receptor and the hypofunctioning of NMDAR has been implicated in the aetiology schizophrenia and specifically impaired learning and memory (29). The schizophrenia-risk allele at p.(Ala391Thr) has also been associated with lower manganese levels (2, 30, 31), an essential trace element transported by ZIP8 and involved in glycosylation, the process by which branched sugar polymers are covalently attached to proteins and lipids (32). Glycosylation is dysregulated in schizophrenia (32) and disrupted by the schizophrenia-risk allele at p.(Ala391Thr) (33). Individuals with Congenital Disorders of Glycosylation can present with cognitive impairment (32), but there is no consistent linear association between lower blood manganese concentrations and poorer cognitive ability (34). Finally, the schizophrenia-risk allele is also associated with impaired zinc uptake and transportation, and decreased cortical dendritic spine density (35). Developmental synaptic pruning has been postulated as a risk mechanism for schizophrenia either through a loss of balance between synaptogenesis and elimination or abnormal activity-dependent plasticity (36). Decreased dendritic spine density has been observed in individuals with schizophrenia (37). Dendritic spine plasticity is thought to underlie the cognitive resilience of older adults who, despite having Alzheimer’s pathophysiology, have not developed dementia (38).

p.(Ala391Thr) has been reported to be a highly pleiotropic variant and is associated, at genome-wide significance levels, with 24.16% (129/534) of the traits curated by Open Targets Genetics (39); the schizophrenia-risk allele has been associated with lower HDL cholesterol, lower blood pressure, lower levels of apolipoprotein A1, calcium, aspartate aminotransferase, urate, gamma-glutamyl transferase, and serum albumin, as well as higher body mass index and body fat measures, but a lower risk of hypertension and cardiovascular disease. Notably, in GWAS, other than in schizophrenia, p.(Ala391Thr) has not been associated with any psychiatric disorder (Bipolar Disorder, Major Depressive Disorder) or with neurodevelopmental (Autism, Attention deficit hyperactivity disorder), or neurodegenerative (Alzheimer’s Disease, Parkinson’s Disease) disorders (see Buniello, MacArthur (40) and Supplementary Table 1). In phenome-wide association studies (PheWAS) using the UK Biobank (UKBB), p.(Ala391Thr) has been associated with diseases of the oesophagus, musculoskeletal conditions, metabolic and digestive biomarkers, blood pressure, and dietary and lifestyle factors including weight gain and drinking alcohol (30, http://www.nealelab.is/uk-biobank/, 41). In a brain MRI PheWAS of the UKBB (42), the schizophrenia risk allele was associated with greater putamen grey matter volume, reduced cortical thickness, and reduced white matter integrity, MRI phenotypes which have been identified in participants with schizophrenia. Although pleiotropy has benefits for translational research, for example by cutting across current diagnostic categories, the diversity of phenotypes suggests that *SLC39A8* may not impact schizophrenia or cognition through the disruption of a single key biological process, rather it may influence multiple processes, not necessarily all within the brain.

As well as increasing our understanding of the aetiology of schizophrenia, studying p.(Ala391Thr) could improve our understanding of the cognitive impairments in people with schizophrenia. Within studies of cognitive remediation therapy (CRT) in schizophrenia, although on average patients with poorer premorbid IQ and fewer years in education show greater improvement after CRT, some studies have shown the opposite (43) and there is emerging literature examining whether genetic variants can account for this differential improvement (44). For example, one study examined *SLC1A2*, a high-affinity glutamate transporter that encodes EAAT2, and found that the minor allele at NC_000011.10:g.35419429T>G (rs4354668), which has previously been associated with lower EAAT2 expression and poorer cognition in healthy controls and patients with schizophrenia, was associated with poorer improvement after CRT (45). p.(Ala391Thr) could be a candidate for future stratification studies.

## Limitations

This is the first study to test the relationship between p.(Ala391Thr) and schizophrenia-related phenotypes in participants with the disorder. However, our cohorts of schizophrenia participants were small, and it may be that larger sample sizes are needed to detect the small effects attributable to a single SNP. Given an alpha level of 0.05, and 90% power, to observe a beta coefficient between -0.07 to -0.02, between N=1741 and N=21403 schizophrenia participants would be required. Our analysis of rare variants in *SLC39A8* may not be representative because of the small number of rare variant carriers driving the associations. Gene-based burden tests usually apply a cut off to exclude genes with a low a number of carriers (46), which, in our sample, would have excluded *SLC39A8* from a pipeline for exome-wide gene-based burden tests. Our rare variant results should therefore be used to inform future studies rather than be interpreted in isolation. It is also possible that our findings will not generalise to participants of non-European ancestry. Previously work has described p.(Ala391Thr) as having an almost negligible minor allele frequency in non-European populations (1). In our schizophrenia cohorts, only Europeans carried two copies of the schizophrenia risk allele but three other ancestry groups (African American/Afro-Caribbean, Near Eastern, and Central/South Asian) contained participants with one copy so we chose to include them in our analysis. Nevertheless, less than 2% of the schizophrenia cohorts were of non-European ancestry and our sample from the UKBB was comprised solely of people with European ancestry.

Another caveat is that it is unclear whether psychotic experiences measured in the UKBB are a good proxy for psychotic experiences experienced by people with schizophrenia. Although there is a shared genetic liability between psychotic experiences and schizophrenia, the genetic correlation is weak (rg = 0.21; 5), and psychotic experiences were more strongly correlated with ADHD (rg = 0.24), autism spectrum disorder (rg = 0.39), and major depressive disorder (rg=0.46) than schizophrenia, suggesting that these phenotypes capture general psychopathology rather than schizophrenia-specific psychosis. There is also the possibility of measurement error in the psychotic experiences phenotypes; firstly, because they are measured retrospectively by self-report, and, secondly, because of biases in those who attempt to complete the online follow-up mental health questionnaire. Legge, Jones (5) found that participants who completed the mental health questionnaire had significantly higher intelligence and lower schizophrenia polygenic risk scores adding to existing evidence of a “healthy volunteer” selection bias in the UKBB (47, 48) which can affect the results of GWAS (49). As the schizophrenia-risk allele was associated with responding to questions in the UKBB with ‘prefer not to answer’ and ‘I don’t know’ (50), we tested whether p.(Ala391Thr) was associated with willingness to attempt the cognitive tests or the mental health questionnaire. p.(Ala391Thr) was not associated with attempting to complete the baseline cognitive tests, but the schizophrenia-risk allele was associated with *not* attempting the mental health questionnaire (see Supplementary Material). This suggests that the psychotic experiences phenotypes in the UKBB are not completed by participants with a higher genetic load for schizophrenia and our analyses may be affected by collider bias.

## Conclusions

The schizophrenia-risk allele at p.(Ala391Thr) is associated with poorer cognitive ability in a sample of the general population with European ancestry. Larger and more ancestrally-diverse studies of participants with schizophrenia are required to determine whether there is an association between p.(Ala391Thr) and cognition in schizophrenia, and subsequently determine whether p.(Ala391Thr) and ZIP8 are potential targets for novel therapeutic treatments for cognitive impairment in those with the disorder.

## Supporting information

Supplementary Material STREGA Checklist

## Data Availability

To comply with the ethical and regulatory framework under which the Cardiff Schizophrenia Samples were obtained, access to individual-level data requires a collaboration agreement with Cardiff University; requests to access these datasets should be directed to J.T.R.W. and M.J.O. UK Biobank data is available by application to the UK Biobank (www.ukbiobank.ac.uk).

## Code Availability

https://zenodo.org/records/10027873

## Acknowledgements

We acknowledge the support of the Supercomputing Wales project, which is part-funded by the European Regional Development Fund (ERDF) via Welsh Government.

## Author Contributions

S.E.S. designed and conducted the analysis and drafted the manuscript. S.E.L. conducted the confirmatory factor analysis. E.F. called the rare variants. A.F.P. conducted the genetic ancestry analysis. A.J.L. scored the MATRICS Consensus Cognitive Battery. V.E-P., E.F., J.H., P.H., S.E.L., M.C.O., M.J.O., A.F.P., G.W., L.W., and J.T.R.W. helped design the analysis and interpret the results. All authors revised and approved the final manuscript.

## Ethical Approval

CardiffCOGS was approved by the South East Wales Research Ethics Committee Panel (reference number: 07/ WSE03/110) and received HRA approval. All participants provided written informed consent. Multicentre and Local Research Ethics Committee approval was obtained for Cardiff F-Series, and all participants gave written informed consent to participate. For Cardiff SibPairs written consent was obtained following local ethical approval guidelines. Ethical approval for the curation and development of DRAGON-Data was obtained from Cardiff University’s School of Medicine Research Ethics Committee (Ref: 19/72). We also used data from the UK Biobank (https://www.ukbiobank.ac.uk), the scientific protocol of which has been reviewed and approved by the North West Multi-centre Ethics Committee. Our access to the UK Biobank data was under the project number 13310.

