## Supplementary Material STREGA Checklist for "SLC39A8.p.(Ala391Thr) is associated with poorer cognitive ability: a cross-sectional study of schizophrenia and the general UK population"

### Reporting checklist for genetic association study.

Based on the STREGA guidelines.

#### Instructions to authors

Complete this checklist by entering the page numbers from your manuscript where readers will find each of the items listed below.

Your article may not currently address all the items on the checklist. Please modify your text to include the missing information. If you are certain that an item does not apply, please write "n/a" and provide a short explanation.

Upload your completed checklist as an extra file when you submit to a journal.

In your methods section, say that you used the STREGAreporting guidelines, and cite them as:

Little J, Higgins JP, Ioannidis JP, Moher D, Gagnon F, von Elm E, Khoury MJ, Cohen B, Davey-Smith G, Grimshaw J, Scheet P, Gwinn M, Williamson RE, Zou GY, Hutchings K, Johnson CY, Tait V, Wiens M, Golding J, van Duijn C, McLaughlin J, Paterson A, Wells G, Fortier I, Freedman M, Zecevic M, King R, Infante-Rivard C, Stewart A, Birkett N; STrengthening the REporting of Genetic Association Studies. STrengthening the REporting of Genetic Association Studies (STREGA): An Extension of the STROBE Statement.

| | | Reporting Item | Page Number |
|---|---|---|---|
| **Title and abstract** | | | |
| Title | #1a | Indicate the study's design with a commonly used term in the title or the abstract | Manuscript – Title (page 2) |
| Abstract | #1b | Provide in the abstract an informative and balanced summary of what was done and what was found | Manuscript – Abstract (page 2) |
| **Background/rationale** | | | |
| | #2 | Explain the scientific background and rationale for the investigation being reported | Manuscript – Introduction (page 3) |

| | | | |
|---|---|---|---|
| **Objectives** | | | |
| | #3 | State specific objectives, including any prespecified hypotheses. State if the study is the first report of a genetic association, a replication effort, or both. | Manuscript – Introduction (page 3) |
| **Study design** | | | |
| | #4 | Present key elements of study design early in the paper | Manuscript – Introduction (page 3) |
| **Setting** | | | |
| | #5 | Describe the setting, locations, and relevant dates, including periods of recruitment, exposure, follow-up, and data collection | Manuscript – Methods (page 3-7)<br><br>Supplementary Methods:<br>• Cardiff Schizophrenia Samples – Participants (page 3)<br>• UK Biobank – Participants (page 7) |
| **Eligibility criteria** | | | |
| | #6a | Cohort study – Give the eligibility criteria, and the sources and methods of selection of participants. Describe methods of follow-up. Case-control study – Give the eligibility criteria, and the sources and methods of case ascertainment and control selection. Give the rationale for the choice of cases and controls. Cross-sectional study – Give the eligibility criteria, and the sources and methods | Manuscript – Methods (page 3-7)<br><br>Supplementary Methods:<br>• Cardiff Schizophrenia Samples – Participants (page 3)<br>• UK Biobank – Participants (page 7) |

| | | | |
|---|---|---|---|
| | | of selection of participants. Give information on the criteria and methods for selection of subsets of participants from a larger study, when relevant. | |
| | #6b | Cohort study – For matched studies, give matching criteria and number of exposed and unexposed. Case-control study – For matched studies, give matching criteria and the number of controls per case. | NA |
| **Variables** | | | |
| | #7a | Clearly define all outcomes, exposures, predictors, potential confounders, and effect modifiers. Give diagnostic criteria, if applicable | Manuscript – Methods (page 3-7)<br><br>Supplementary Methods:<br>• Cardiff Schizophrenia Samples – Phenotypes (page 3-4)<br>• UK Biobank – Phenotypes (page 7-11) |
| | #7b | Clearly define genetic exposures (genetic variants) using a widely-used nomenclature system. Identify variables likely to be associated with population stratification (confounding by ethnic origin). | Manuscript – Methods (page 3-7)<br><br>Supplementary Methods:<br>• Cardiff Schizophrenia Samples – Genotypes (page 4-6)<br>• UK Biobank – Genotypes (page 11-12) |
| **Data sources/measurement** | | | |
| | #8a | For each variable of interest give sources of data and details of methods of assessment (measurement). Describe comparability of | Manuscript – Methods (page 3-7)<br><br>Supplementary Methods: |

| | | | |
|---|---|---|---|
| | | assessment methods if there is more than one group. Give information separately for for exposed and unexposed groups if applicable. | • Cardiff Schizophrenia Samples – Phenotypes (page 3-4)<br>• UK Biobank – Phenotypes (page 7-11) |
| | #8b | Describe laboratory methods, including source and storage of DNA, genotyping methods and platforms (including the allele calling algorithm used, and its version), error rates and call rates. State the laboratory / centre where genotyping was done. Describe comparability of laboratory methods if there is more than one group. Specify whether genotypes were assigned using all of the data from the study simultaneously or in smaller batches. | Supplementary Methods:<br>• Cardiff Schizophrenia Samples – Genotypes (page 4-6)<br>• UK Biobank – Genotypes (page 11-12) |
| **Bias** | | | |
| | #9a | Describe any efforts to address potential sources of bias | Supplementary Methods:<br>• Cardiff Schizophrenia Samples – Genotypes – Genetic ancestry (page 5-6)<br>• UK Biobank – Participants – Attempted phenotypes of interest (page 7) |
| | #9b | Describe any efforts to address potential sources of bias | NA |
| **Study size** | | | |
| | #10 | Explain how the study size was arrived at | Manuscript – Methods (page 3-7) |

| | | | |
|---|---|---|---|
| | | | Supplementary Methods:<br>• Cardiff Schizophrenia Samples – Participants (page 3)<br>• Cardiff Schizophrenia Samples – Genotypes (page 4-6)<br>• UK Biobank – Participants (page 7)<br>• UK Biobank – Genotypes (page 11-12) |
| **Quantitative variables** | | | |
| | #11 | Explain how quantitative variables were handled in the analyses. If applicable, describe which groupings were chosen, and why. If applicable, describe how effects of treatment were dealt with. | Manuscript – Methods (page 3-7)<br><br>Supplementary Methods:<br>• Cardiff Schizophrenia Samples – Phenotypes (page 3-4)<br>• UK Biobank – Phenotypes (page 7-11)<br>• Statistical Analysis (page 12) |
| **Statistical methods** | | | |
| | #12a | Describe all statistical methods, including those used to control for confounding. State software version used and options (or settings) chosen. | Manuscript – Methods – Statistical Analysis (page 7)<br><br>Supplementary Methods:<br>• Statistical Analysis (page 12) |
| | #12b | Describe any methods used to examine subgroups and interactions | NA |
| | #12c | Explain how missing data were addressed | Manuscript – Methods – Statistical Analysis (page 7)<br><br>Supplementary Methods: |

| | | |
|---|---|---|
| | | Statistical Analysis (page 12) |
| #12d | If applicable, explain how loss to follow-up was addressed | NA |
| #12e | Describe any sensitivity analyses | NA |
| #12f | State whether Hardy-Weinberg equilibrium was considered and, if so, how. | Supplementary Methods:<br>• Cardiff Schizophrenia Samples – Genotypes (page 4-6)<br>• UK Biobank – Genotypes (page 11-12) |
| #12g | Describe any methods used for inferring genotypes or haplotypes | Supplementary Methods:<br>• Cardiff Schizophrenia Samples – Genotypes – Imputation (page 5)<br>• UK Biobank – Genotypes – Imputation (page 11) |
| #12h | Describe any methods used to assess or address population stratification. | Supplementary Methods:<br>• Cardiff Schizophrenia Samples – Genotypes – Principal components (page 6)<br>• UK Biobank – Genotypes – Population stratification (page 11) |
| #12i | Describe any methods used to address multiple comparisons or to control risk of false positive findings. | Manuscript – Methods – Statistical Analysis (page 7) |
| #12j | Describe any methods used to address and correct for relatedness among subjects | Supplementary Methods:<br>• Cardiff Schizophrenia Samples – Genotypes – Post-imputation QC for this study (page 5)<br>• UK Biobank – Genotypes – Post-imputation QC for this study (page 11) |

**Participants**

| | | | |
|---|---|---|---|
| | #13a | Report numbers of individuals at each stage of study—eg numbers potentially eligible, examined for eligibility, confirmed eligible, included in the study, completing follow-up, and analysed. Give information separately for for exposed and unexposed groups if applicable. Report numbers of individuals in whom genotyping was attempted and numbers of individuals in whom genotyping was successful. | See previous studies cited in<br><br>Supplementary Methods:<br>• Cardiff Schizophrenia Samples – Participants (page 3)<br>• UK Biobank – Participants (page 7) |
| | #13b | Give reasons for non-participation at each stage | Unknown |
| | #13c | Consider use of a flow diagram | NA |

**Descriptive data**

| | | | |
|---|---|---|---|
| | #14a | Give characteristics of study participants (eg demographic, clinical, social) and information on exposures and potential confounders. Give information separately for exposed and unexposed groups if applicable. Consider giving information by genotype | Manuscript – Results (page 8)<br><br>Supplementary Results:<br>• Cardiff Schizophrenia Samples – Descriptive statistics (page 13)<br>• UK Biobank – Descriptive statistics (page 14)<br><br>Supplementary Tables:<br>• Table 5<br>• Table 6<br>• Table 7<br>• Table 22 |

| | | | |
|---|---|---|---|
| | #14b | Indicate number of participants with missing data for each variable of interest | Supplementary Tables:<br>• Table 4<br>• Table 21 |
| | #14c | Cohort study – Summarize follow-up time, e.g. average and total amount. | NA |
| **Outcome data** | | | |
| | #15 | Cohort study Report numbers of outcome events or summary measures over time.Give information separately for exposed and unexposed groups if applicable. Report outcomes (phenotypes) for each genotype category over time Case-control study – Report numbers in each exposure category, or summary measures of exposure.Give information separately for cases and controls . Report numbers in each genotype category. Cross-sectional study – Report numbers of outcome events or summary measures. Give information separately for exposed and unexposed groups if applicable. Report outcomes (phenotypes) for each genotype category | Supplementary Results:<br>• Cardiff Schizophrenia Samples – Descriptive statistics (page 13)<br>• UK Biobank – Descriptive statistics (page 14)<br><br>Supplementary Tables:<br>• Table 5<br>• Table 6<br>• Table 7<br>• Table 22 |
| **Main results** | | | |
| | #16a | Give unadjusted estimates and, if applicable, confounder-adjusted estimates and their precision (eg, 95% confidence | SM Supplementary Tables:<br>• Table 8<br>• Table 9<br>• Table 10<br>• Table 23 |

| | | | |
|---|---|---|---|
| | | interval). Make clear which confounders were adjusted for and why they were included | • Table 24 |
| | #16b | Report category boundaries when continuous variables were categorized | NA |
| | #16c | If relevant, consider translating estimates of relative risk into absolute risk for a meaningful time period | NA |
| | #16d | Report results of any adjustments for multiple comparisons | Manuscript – Results (page 8)<br><br>SM Supplementary Tables:<br>• Table 8<br>• Table 9<br>• Table 10<br>• Table 23<br>• Table 24 |
| **Other analyses** | | | |
| | #17a | Report other analyses done—e.g., analyses of subgroups and interactions, and sensitivity analyses | NA |
| | #17b | Report other analyses done—e.g., analyses of subgroups and interactions, and sensitivity analyses | NA |
| | #17c | Report other analyses done—e.g., analyses of subgroups and interactions, and sensitivity analyses | NA |
| **Key results** | | | |
| | #18 | Summarise key results with reference to study objectives | Manuscript – Discussion (page 9) |

**Limitations**

| | | | |
|---|---|---|---|
| | #19 | Discuss limitations of the study, taking into account sources of potential bias or imprecision. Discuss both direction and magnitude of any potential bias. | Manuscript – Discussion – Limitations (page 10-11) |

**Interpretation**

| | | | |
|---|---|---|---|
| | #20 | Give a cautious overall interpretation considering objectives, limitations, multiplicity of analyses, results from similar studies, and other relevant evidence. | Manuscript – Discussion (page 9-11) |

**Generalisability**

| | | | |
|---|---|---|---|
| | #21 | Discuss the generalisability (external validity) of the study results | Manuscript – Discussion (page 9-11) |

**Funding**

| | | | |
|---|---|---|---|
| | #22 | Give the source of funding and the role of the funders for the present study and, if applicable, for the original study on which the present article is based | Manuscript – Funding (page 1) |

None The STREGA checklist is distributed under the terms of the Creative Commons Attribution License CC-BY. This checklist can be completed online using https://www.goodreports.org/, a tool made by the EQUATOR Network in collaboration with Penelope.ai
