## Supplementary Material Text for "SLC39A8.p.(Ala391Thr) is associated with poorer cognitive ability: a cross-sectional study of schizophrenia and the general UK population"

### Supplementary Methods

#### Cardiff Schizophrenia Samples

##### *1. Participants*

Data from CardiffCOGS has been previously reported (1-4); as has data from Cardiff F-Series (5) and Cardiff SibPairs (6-8).

###### *1.1. Inclusion/exclusion criteria*

In CardiffCOGS, participants were excluded if they had a neurologic condition that was likely to affect their ability to participate in the study, or if they had a current substance dependence disorder. Inter-rater reliability for research diagnosis has been previously described (2). Participants were interviewed using the Schedules for Clinical Assessment in Neuropsychiatry (SCAN) (9), a semi-structured interview, which was then reviewed by trained raters who, along with available clinical records, determined a consensus lifetime DSM-IV diagnosis (inter-rater reliability κ statistics: schizophrenia = 0.83, schizoaffective depressed type = 0.63).

In Cardiff F-Series and Cardiff SibPairs, participants were interviewed using the SCAN or Present State Examination (PSE) (10), which, along with available clinical records, was used by raters to make a consensus diagnoses (5, 8).

##### *2. Phenotypes*

###### *2.1. Symptoms dimensions*

###### *2.1.1. SAPS and SANS*

The Scale for the Assessment of Positive Symptoms (SAPS) (11) and the Scale for the Assessment of Negative Symptoms (SANS) (12) were scored on a lifetime worst basis using information from the SCAN interview and lifetime psychiatric clinical records. For each participant, four global domain scores were calculated using the SAPS (bizarre behaviour, delusions, hallucinations, and positive thought disorder), four global domain scores were calculated using the SANS (affective flattening, alogia, anhedonia/asociality, and avolition/apathy), and one individual item from the SANS (inappropriate affect) was scored. Global domain scores were scored on an ordinal scale (0-5). Inter-rater reliability for the SAPS and SANS ratings was good, ranging from 0.72 to 0.95 across the three samples (13).

###### *2.1.2. MATRICS Consensus Cognitive Battery*

As described in Lynham, Hubbard (2), the MATRICS Consensus Cognitive Battery measures 7 domains of cognition using 10 tasks: (1) speed of processing (Brief Assessment of Cognition in Schizophrenia: Symbol Coding; Category Fluency: Animal Naming; Trail Making Test: Part A); (2) working memory (Wechsler Memory Scale III: Spatial Span; Letter-Number Span); (3) attention/vigilance (Continuous Performance Test: Identical Pairs); (4) verbal learning (Hopkins Verbal Learning Test–Revised); (5) visual learning (Brief Visuospatial Memory Test–Revised); (6) Reasoning and problem-solving (Neuropsychological Assessment Battery: Mazes); and (7) social cognition (Mayer-Salovey-Caruso Emotional Intelligence Test: Managing Emotions). For each task, z scores were derived using the mean and standard deviation of a control group. Controls (N=103, 50% men, mean age 41.7 years) completed the Mini International Neuropsychiatric Interview (Sheehan et al., 1998) as a screen for mental disorders and were excluded if they met criteria for schizophrenia or bipolar disorder or had a family history of these conditions. For each participant, MATRICS Consensus Cognitive Battery domain scores were calculated for the seven domains, following the MCCB manual procedures, and scored on a continuous scale.

##### 2.1.3. *Confirmatory factor analysis*

Symptom dimensions are considered more appropriate than rating scale scores for studies of disorders where not all participants exhibit the same symptoms (14). As described in Legge, Cardno (13) a confirmatory factor analysis (CFA) framework was used to estimate phenotype-derived dimension scores. Fit was assessed using comparative fit index (CFI; of at least 0.95), root mean square error of approximation (RMSEA; values lower than 0.06), and standardized root mean square residual (SRMR; lower than 0.08). Using all three samples, the best model had three symptom dimensions: positive symptoms (SAPS hallucinations and SAPS delusions), negative symptoms of diminished expressivity (SANS affective flattening and SANS alogia), and disorganised symptoms (SAPS positive formal thought disorder and SANS inappropriate affect). This model was a good fit for the data: CardiffCOGS, CFI=0.99, RMSEA=0.05 (95%CI 0.02-0.07), SRMR=0.04; Cardiff F-series, CFI=1.00, RMSEA=0.00 (95%CI 0.00-0.05), SRMR=0.04; and Cardiff Affected-Sib, CFI=0.99, RMSEA=0.05 (95%CI 0.00-0.00), SRMR=0.04. Using only the CardiffCOGS sample, a second CFA framework was used to estimate phenotype-derived factor scores from the symptom ratings and the MATRICS consensus cognitive battery (15). The best model had five symptom dimensions: positive symptoms (as above), negative symptoms of diminished expressivity (as above), disorganised symptoms (as above), negative symptoms of motivation and pleasure (SANS anhedonia/asociality and SANS avolition/apathy), and cognitive ability (all MATRICS domains except social cognition). This model was a good fit for the data: CFI=0.99, RMSEA=0.04 (95%CI 0.03-0.05), SRMR=0.05.

#### 2.2. *Premorbid IQ*

The National Adult Reading Test (NART) (16), where individuals are required to read and pronounce a list of 50 words, was standardised within the Cardiff Schizophrenia Samples.

#### 2.3. *Educational attainment*

In all three samples, a participant's highest educational attainment was recorded using six categories: (1) none; (2) eleven-plus (also known as 11+); (3) Certificate of Secondary Education (CSE); (4) General Certificate of Education: Ordinary Level (also known as Ordinary Level or O Level)/General Certificate of Secondary Education (GCSE); (5) General Certificate of Education: Advanced Level (also known as A Level)/Higher National Diploma (HND)/Business and Technology Education Council (BTEC) qualification/Diploma (otherwise unspecified)/Certificate (otherwise unspecified); and (6) Degree. We derived two binary variables: 'GCSE' vs. 'No GCSE' [4 vs <4 according to coding] and 'Degree' vs. 'No Degree' [6 vs <6 according to coding].

#### 2.4. *Age of psychosis onset*

As well as the symptom domains, psychosis illness severity was assessed using age at psychosis onset. Younger age of psychosis onset has been associated with family history of psychosis (17), increased rates of hospitalisation and relapse (18), poorer social and occupational functioning (18), and antipsychotic treatment resistance (19).

### 3. *Genotypes*

The genetic data for the Cardiff Schizophrenia Samples was curated and harmonised with genetic information from multiple cohorts within the MRC Centre for Neuropsychiatric Genetics and Genomics to create a new data repository, DRAGON-DATA. Details of the genetic quality control (QC) have been fully described in Lynham, Knott (20) and are summarised below.

#### 3.1. *Genotyping*

Participants were genotyped on the Illumina OmniExpress (Infinium OmniExpress-24 Kit) genotyping chip (version 8 or version 12). SNP IDs were annotated using Genome Reference Consortium Human Build 37 (GRCh37/hg19).

#### 3.2. Pre-imputation QC

Pre-imputation QC was performed using Plink v1.9 (21). SNPs were aligned to the haplotype reference consortium (HRC) (22) using Genotype Harmonizer (23). QC was performed separately on each study in the DRAGON_DATA. Participants were removed if (1) their genetic sex was ambiguous were removed (male sex was defined as F ≥ 0.8 and female sex is defined as F ≤ 0.2), (2) their genetic sex did not match their self-reported gender, and (3) if they were missing more than 5% of the available single nucleotide polymorphism (SNP). Next, SNPs were removed if (1) they had a minor allele frequency (MAF) less than 1%, (2) they were missing for more than 5% of individuals, (3) they were in deviation from Hardy-Weinberg Equilibrium (([using the *midp* and *keep-fewhet* flags in Plink2] ≤ $10^{-6}$). Using this QC'd sample, duplicates were identified using kinship coefficient greater than 0.9 and removed (known identical twins were not removed) and relatives were identified but not removed. Cohorts genotyped on the same platform were grouped into batches based on their genotyping platform and the above QC steps were repeated for each batch separately.

#### 3.3. Imputation

Each batch was imputed separately. Genotypes were uploaded to the Michigan Imputation Server (24) and missing genotypes imputed using the Haplotype Reference Consortium V1.1. reference panel (22) (Eagle v2.4 for phasing, Minimac4). Along with all the DRAGON-DATA cohorts, large datasets of healthy controls were also uploaded in the same batches: Gen Scotland and POBI for the OmniExpress batch.

#### 3.4. Post-imputation DRAGON-Data QC

Post-imputation QC was performed using Plink v2. SNPs were removed if (1) they have a individual genotype probability threshold less than 0.9, (2) they had a MAF less than 1%, (3) they were missing for more than 5% of individuals, (4) they were in deviation from Hardy-Weinberg Equilibrium (mid-p ≤ $10^{-4}$) (25). Dosage data was then converted to best guess genotype data and SNPs with an imputation quality INFO score less then 0.3 were removed.

#### 3.5. Post-imputation QC for this study

This sample was then restricted to participants with an ICD or DSM diagnosis of schizophrenia or schizoaffective disorder, depressed type. SNPs were restricted to those with an INFO score > 0.9. First-degree relatives (pi-hat estimate of identity-by-descent; $\pi > 0.4$), both within and between samples, had previously been identified and so we removed one member from each related pair, preferentially retaining samples that had more complete phenotype data and otherwise removed at random (13). We further removed related individuals, including relatives' half-way between second and third degree, using the KING-robust method which can infer relatedness in the presence of population substructure (26). Relationship inference criteria are based on estimating kinship coefficients ($\varphi$) and the probability of zero IBD sharing ($\pi_0$). We used a cutoff of 0.09375, the geometric mean between second-degree relatives (0.125) and third-degree relatives (0.0625). We then removed SNPs if (1) they had a MAF less than 1%, (2) they were missing for more than 5% of individuals, (3) they were in deviation from Hardy-Weinberg Equilibrium (mid-p keep-fewhet ≤ $10^{-6}$) (25).

#### 3.6. Genetic ancestry

Using pre-imputation QC'd genotypes, for each study separately, global ancestry inference for all samples was performed using the linear discriminant analysis (LDA) method described in Legge, Pardiñas (27) with the following modifications. First, to increase the robustness of the algorithm we replaced the original Human Genome Diversity Project (HGDP) reference panel (N=930; 28, 29) with a larger and more diverse sample from the Allen Ancient DNA Resource (AADR) (https://reich.hms.harvard.edu/allen-ancient-dna-resource-aadr-downloadable-genotypes-present-day-and-ancient-dna-data; v50), after restricting to contemporaneous individuals and removing sample duplicates (N=6096). Second, for consistency with recently published literature, we applied the biogeographical groups defined by Huddart, Fohner (30) to the AADR sample and used them as training classes for the algorithm; seven biogeographical categories: American, East Asian, European, Central/South Asian, Near Eastern, Oceanian, Sub-Saharan African, and two admixed categories: African American/Afro-Caribbean and Latino. For a graphical representation of these biogeographical regions please see Figure 1 in Huddart, Fohner (30). Third, the best probability threshold for determining an ancestral category was not defined as a fixed value, but determined within each ancestry using Youden's index as optimality criterion via the "probably" R package (https://probably.tidymodels.org/). Other than this, all other aspects of the ancestry inference pipeline (AIM definition and PCA-based dimensionality reduction) were performed following the original publication.

To define the Ancestry Informative Markers (AIMs) we used the $F_{ST}$-based procedure of Kersbergen, van Duijn (31), which requires defining the desired ancestry groups a priori. Thus, we divided the AADR sample into the biogeographical categories (30), and calculated pairwise between-group $F_{ST}$ metrics for each SNP using PLINK v1.9. We retained all pairwise comparisons. For each of these, we selected all the SNPs in the top 2.5 percentile of the $F_{ST}$ values (29), assuming that this metric follows a beta distribution (32). The resulting SNPs were then pruned ($r^2$=0.4) using the linkage disequilibrium (LD) structure of their corresponding non-European population (i.e. the SNPs selected from the Europe-Africa pairwise comparison were LD-pruned using the African samples as reference). In order to reduce the AIMs to a small set of independent variables we used the principal component analysis (PCA) implemented in EIGENSTRAT v6.12 (33) on the AADR AIM SNPs. Then, to retain a number of principal components proportional to the level of population structure in the sample, we used a Tracy-Widom test (34). In order to classify the Cardiff individuals we merged the study genotype data with the AADR AIMs and repeated the generation of PCs using the EIGENSTRAT “projection” mode, as described above.

In summary, the global LDA model used in the CardiffCOGS sample was based on 29,225 AIMs collapsed to 43 PCs. Balanced accuracies (sens+spec/2) (35) for ancestry classification of these LDA models ranged between 88.54% and 99.97% as estimated by 10-fold cross-validation. The F-Series and Sibpairs samples were processed together and the global LDA model used 30,411 AIMs collapsed to 41 PCs. The accuracy range on training data for this model was 91.41%-99.97%.

Within-European ancestry inference was again determined as above but using a smaller European-only AADR reference panel (N=1998) in which the training classes were defined as the European subregions labelled in the AADR metadata file. The European LDA model used in the CardiffCOGS sample was based on 3,264 AIMs collapsed to 19 PCs. Balanced accuracies for ancestry classification of these LDA models ranged between 89.43% and 97.32% as estimated by 10-fold cross-validation. The European LDA model used in the F-Series and Sibpairs samples was based on 10,288 AIMs collapsed to 17 PCs. The accuracy range on training data for this model was 92.17%-98.63%.

###### *3.6.1. Association between rs13107325 and genetic ancestry*

The schizophrenia allele for rs13107325 was only present in four ancestry categories (Table 2), participants within these groups were retained for the analyses. No ancestry probabilities were associated with rs13107325 allele count at alpha level < 0.05 (Table 3).

##### *3.7. Principal components*

After the data had been restricted to ancestries where the schizophrenia-risk allele was present, we removed 24 autosomal long-range LD regions, as identified by Price, Weale (36). These regions, on human genome build GRCh37/hg19, were: 1:48–52 Mb, 2:86–100.5 Mb, 2:134.5–138 Mb, 2:183–190 Mb, 3:47.5–50 Mb, 3:83.5–87 Mb, 3:89–97.5 Mb, 5:44.5–50.5 Mb, 5:98–100.5 Mb, 5:129–132 Mb, 5:135.5–138.5 Mb, 6:25–35 Mb, 6:57–64 Mb, 6:140–142.5 Mb, 7:55–66 Mb, 8:7–13 Mb, 8:43–50 Mb, 8:112–115 Mb, 10:37–43 Mb, 11:46–57 Mb, 11:87.5–90.5 Mb, 12:33–40 Mb, 12:109.5–112 Mb, and 20:32–34.5 Mb. Next, we calculated principal components to identify and account for population and ancestral substructure. We used independent pairwise Linkage Disequilibrium-based to prune the SNPs (500 variant count window size, 1 variant count to shift the window at the end of each step, a pairwise $r^2$ threshold of 0.2). Plink 2.0 was used to calculate principal components using these pruned SNPs. PCs1-5 were used as covariates in all analyses. In addition, PCs 6 and 9 were associated with the schizophrenia-risk allele in the Cardiff F-Series sample and so were also included these as covariates when analysing this sample.

#### UK Biobank

##### 1. *Participants*

###### *1.1. Psychotic spectrum disorder*

All individuals in the UKBB have their health records linked so we removed participants with a psychotic spectrum disorder. Participants with a psychotic spectrum disorder (F20 schizophrenia [UKBB Fields: 130875, 20002, 40001, 40002, 41202, 41204, 20544], F21 schizotypal disorder [UKBB Fields: 130877], F22 persistent delusional disorders [UKBB Fields: 130879], F23 acute and transient psychotic disorders [UKBB Fields: 130881], F24 induced delusional disorders [UKBB Fields: 130883], F25 schizoaffective disorder [UKBB Fields: 130885], F28 other nonorganic psychotic disorders [UKBB Fields: 130887], F29 unspecified nonorganic psychosis [UKBB Fields: 130889]) were identified from the hospital, death, and primary care records (ICD-10 codes), as well through self-report at the assessment centre interview and in the online follow-up mental health questionnaire (see https://biobank.ndph.ox.ac.uk/showcase/ukb/docs/mental_health_online.pdf).

###### *1.2. Withdrawn participants*

We excluded participants who had requested their data be withdrawn from the UKBB up to 25/04/2023.

###### *1.3. Attempted phenotypes of interest*

We further restricted the sample to participants who'd indicated that they were willing to attempt the cognitive tests at baseline [UKBB Fields: 62] or attempted to complete the online follow-up mental health questionnaire [non-missing data for 'date of completing mental health questionnaire'; UKBB Fields: 20400].

Given that specific genetic loci have been found to be associated with questionnaire response behaviour using the UKBB (37), we tested whether rs13107325 was associated with attempting to complete the baseline cognitive tests [UKBB Fields: 62] and attempting to complete the online follow-up mental health questionnaire [non-missing data for date of completing mental health questionnaire; UKBB Fields: 20400]. We used the subsample of UKBB participants who self-reported European ancestry [UKBB Fields: 22006] after excluding those with a psychotic spectrum disorder or who requested their data be withdrawn. After adjusting for age at assessment, age at interview squared, sex, the first ten genetic PCs, and genotype batch, rs13107325 was not associated with attempting or not attempted to complete the baseline cognitive tests (N=401303 attempted, N=4906 did not attempt, OR = 1.06; 95% CI, 0.98-1.14; p-value = 0.128) but the schizophrenia-risk allele was associated with *not* attempting the online follow-up mental health questionnaire (N=131442 attempted, N=275142 did not attempt, OR = 0.97; 95% CI, 0.95-0.99; p-value = 0.001).

###### *1.4. Ancestry*

Finally, we restricted the sample to participants of European ancestry, as defined by the UKBB [UKBB Fields: 22006] and then to a genetically homogenous subgroup (see 3.4.1).

##### 2. *Phenotypes*

###### *2.1. Psychotic experiences*

Data on psychotic experiences were collected using questions in Section F of the online follow-up mental health questionnaire. As of July 2017, 339,229 participants had been sent an email invitation to complete the mental health questionnaire, of whom 158,835 (46.8%) fully completed the questionnaire. A further 416 participants accessed the questionnaire via the participant website without having received an email invite (because they have not provided UK Biobank with a valid email address).

Section F was designed to assess unusual experiences that may be markers of a tendency towards psychosis or may be a harbinger of neurodegenerative disease and is based on the World Health Organization Composite International Diagnostic Interview (CIDI) psychosis module (lifetime version) (38). The CIDI is a structured diagnostic interview designed to assess the prevalence of DSM-IV and ICD-10 mental disorders. The original CIDI psychosis module includes questions about six psychotic experiences: two hallucinatory experiences and two bizarre delusionary experiences and two paranoid delusionary experiences. The UKBB mental health questionnaire is an abridged version of the CIDI and includes questions (see Table 14) from the psychosis module about two hallucinatory experiences (visual [20463] and auditory [20471] hallucinations) and two paranoid delusionary experiences (ideas of reference [20474] and plot to harm and/or follow i.e. delusions of persecution [20468]).

As in Legge, Jones (39), we derived three overlapping binary variables: (1) any psychotic experience defined as a positive response to any of the four symptom questions [UKBB Fields: 20463, 20468, 20471, and 20474]; (2) a distressing psychotic experience, defined as any psychotic experience that was rated as “a bit,” “quite,” or “very” distressing [UKBB Fields: 20462]; and (3) multiple occurrences of psychotic experiences, defined as any psychotic experience that occurred on more than 1 occasion [UKBB Fields: 20465, 20470, 20473, and 20476]. The comparator group for these three variables was comprised of individuals who provided a negative response to all four psychotic experience symptom questions. We also looked at delusions of persecution [20468] alone as previous work has suggested that this phenotype in the UKBB is particularly enriched for genetic liability for schizophrenia (39).

##### 2.2. Cognitive ability

The cognitive assessment was administered as part of the fully-automated touchscreen questionnaire (40). The original UK Biobank cognitive assessment was very brief (approximately 5 minutes). At baseline (A1, initial assessment visit [2006-2010]), almost all ~500,000 participants completed tests of pairs matching (visual memory) and reaction time (processing speed). Sub-samples also completed tests of numeric memory (working memory) and fluid intelligence (verbal and numerical reasoning) (see https://biobank.ndph.ox.ac.uk/showcase/ukb/docs/Orderofdatacollection.pdf for order of data collection). This test battery, excluding numeric memory and including a matrix pattern completion test (reasoning), was then administered again (A2, first repeat assessment visit [2012-2013]) where ~20,000 participants were re-tested. This test battery was then administered again at two further assessment visits (A3, imaging visit [2014+] and A4, first repeat imaging visit [2019+]). Additional tests of symbol digit substitution (processing speed), tower rearranging (executive function), and trail making tests (processing speed and executive function) were included. In addition to these assessment centre-based visits, subsamples have completed online cognitive tests (including, in the following order, fluid intelligence, trail making, symbol digit substitution, pairs matching, and numeric memory). A study in an independent sample found that the tests used in the UKBB had moderate-to-high test-retest reliability (41). Each individual test is described in detail below.

Across all tests which were conducted at multiple testing centres and/or online, only one score (from the first time a participant completed the test) was used for each participant. Assessment centres in order of date were prioritised, and the online assessment was used only if an individual had not previously completed the test at any in-person study visit. For each test, we used the assessment which had the most participants completing the test for the first time (Table 15). The normality of raw scores was assessed through visual inspection of histograms and the calculation of skewness and kurtosis. If required, a log(1+x) transformation was performed (Table 16). For each test, participants’ raw scores were converted into z-scores using the mean and standard deviation of the sample with non-missing data for that test. Participants with a z-score ≥ 4 or ≤ 4 standard deviations from the mean were identified as outliers and excluded.

###### 2.2.1. Fluid intelligence

We chose to use fluid intelligence as a measure of cognition on its own, therefore, it was not considered for inclusion in the g score [see below]). The fluid intelligence test is often used as a measure of current cognitive ability in the literature and could therefore be used as a complementary measure of cognition.

##### 2.2.2. *g score*

Performance on different cognitive tests is highly correlated, both in the UKBB (41, 42) and in other samples (43, 44). As a consequence, cognitive test scores, spanning multiple domains, can be used to calculate a general intelligence factor, g, which is a considered a measure of cognitive ability. As in Fawns-Ritchie and Deary (41) and Lyall, Cullen (42), we calculated g using principal component analysis (PCA) of several cognitive tests. The first unrotated principal component from a PCA of cognitive tests was defined as g. As there was no way to covary for the source of the test score (e.g. whether it was completed at an assessment centre or online), only one occurrence of each test was used to form g. In every case, this was the occurrence with the highest number of participants from Table 15. g could only be calculated for participants with complete data on the tests which went into the PCA.

We selected four cognitive tests to form g (numeric memory (online), reaction time, pairs matching, and trail making test (TMT) B). These test were chosen because (1) they maximised the number of participants for which g could be calculated (tests with low completion rates (<20% across all assessments) were not considered for inclusion i.e. matrix pattern completion, tower rearranging) (see Table 15 and Figure 1), (2) they were not highly correlated with one another (Table 17), and (3) they each measured a different cognitive domain allowing g to represented broad cognition (Table 15), i.e. reaction time, symbol digit substitution, and trail making test A all measure processing speed but reaction time had the largest sample size. Numeric memory (online) was reversed so that, for all four tests, lower scores represented better cognitive ability.

g was then standardised by converting values to z scores, outliers above and below 4 standard deviations of the mean were removed, and z-scores were multiplied by -1 to enable a more intuitive reading of g; a positive g score represents better cognitive performance. g was calculated for N = 54858 participants, accounted for 38.80% of the variance in the tests (Table 18), and was well correlated with both the tests that formed g and those which did not, to greater extent than the fluid intelligence score (Table 19 and Figure 2).

##### 2.2.3. *Descriptions of each individual cognitive test*

Fluid intelligence (FI)

This test of reasoning presented participants with multiple-choice questions to measure their problem solving, logic, and reasoning capacity independent of acquired knowledge. They were given two minutes to answer as many multiple-choice questions as possible (with a maximum of 13) and the questions got progressively harder. The number of correct answers given was used as the outcome measure [UKBB Fields: 20016 and 20191]. For the online assessment, participants who abandoned the test were excluded [UKBB Field: 20242]. Participants who did not answer all of the questions within the allotted two-minute limit were scored as zero for each of the unattempted questions. The initial question involved selecting the largest number out of five three-digit numbers presented and the thirteenth question asked participants to select whether the following statement is true, false, or neither: "If some flinks are plinks and some plinks are stinks then some flinks are definitely stinks?". The first study visit had the largest number of participants who were completing the test for the first time.

Matrix Pattern Completion (MPC)

This tested reasoning and involved the presentation of a series of blocks forming a pattern, with one element missing. The participant was asked to select which element of a number of options best completes the pattern. The number of puzzles correctly solved (maximum total score 15) was used as the outcome measure for this test (UKBB Field 6373). Participants who did not view all 15 puzzles were assumed to have abandoned the test and excluded [UKBB Field: 6374]. The second study visit had the largest number of participants who were completing the test for the first time.

Numeric Memory (NM)

This tested working memory and involved asking participants to remember numbers, which progressively increased by one digit, and input them immediately after the presentation. The outcome measure we used was the number of digits correctly remembered [UKBB Fields: 4284 and 20240]. Participants who were coded as abandoned the test [-1] were excluded. The online assessment had the largest number of participants who were completing the test for the first time.

Pairs Matching (PM)

This test of declarative memory was completed by almost all participants. A matrix of face-up cards was presented, and participants were asked to remember the position of as many pairs of matching cards as possible. Cards were then turned face-down, and participants selected the pairs of cards. Two rounds of this task were run, with the second involving a larger number of cards, and therefore pairs, to remember. The summed total of the number of incorrect matches across these two rounds was used as the outcome measure [UKBB Fields: 399 and 20132]. For the online assessment, participants who abandoned the test were excluded [UKBB Field: 20244]. The first study visit had the largest number of participants who were completing the test for the first time.

Reaction Time (RT)

This test of processing speed was very similar to a game of 'snap', played across 12 rounds. Almost all participants completed this test. The participant was shown two cards and asked to press a button if they matched. The mean time taken to press the button, in rounds in which both cards matched, was used as the outcome measure [UKBB Field 20023]. The following rounds were not used to calculate this mean: the first four training rounds which were considered training rounds; rounds with times under 50ms which captured anticipation rather than reaction; and rounds over 2000ms, the time at which the cards disappeared. Values were rounded to the nearest whole number. The first study visit had the largest number of participants who were completing the test for the first time.

Symbol Digit Substitution (SDS)

This test of processing speed involved presenting a key of symbols, matched to numbers, to the participant and asking the participant to indicate which number matched each symbol in a grid. The number of correct matches was used as the outcome measure [UKBB Fields: 23324 and 20159]. For the online assessment, participants who abandoned the test were excluded [UKBB Field: 20245]. The online assessment had the largest number of participants who were completing the test for the first time.

Tower Re-arranging (TR)

This test of executive function asked participants to determine the minimum number of moves required to re-arrange hoops on three pegs (towers) from one position to another given position. The number of puzzles correctly solved was used as the outcome measure [UKBB Field: 21004]. The third study visit had the largest number of participants who were completing the test for the first time.

Trail Making Tests (TMT)

The trail making tests measured executive function and processing speed. For TMT A, participants were presented with a set of digits scattered across the screen and asked to click on them in numeric order. TMT B was similar, but both letters and digits were presented and participants were asked to click on them in alphanumeric order (e.g. A-1-B-2-C … ). For both tests, the outcome measure was the time taken to complete the trail [UKBB Fields: 20156 and 20157]. The online assessment had the largest number of participants who were completing the both tests for the first time.

##### 2.3. *Educational attainment*

In the UKBB, participants' educational qualifications were recorded using six non-exclusive categories [UKBB Field: 6138] (Table 20). We used two binary variables: 'GCSE' vs. 'No GCSE' [3 vs >3 according to UKBB coding] and 'Degree' vs. 'No Degree' [1 vs >1 according to UKBB coding]. Note that if someone misread the question and only selected their highest educational qualification, and this was higher than a GSCE, they will not be included in the binary variable for GCSE.

Educational attainment was also measured using years in education [UKBB Field: 845], but only for those without a college or university degree. Those who selected "Never went to school", "Do not know", or "Prefer not to answer" were coded as missing.

##### 2.4. Age squared

Age squared was included as a covariate in the regression models when a measure of cognitive ability was used as the outcome. Non-linear relationships between age and the UKBB cognitive test have been observed (45). Since age squared is a polynomial term derived from age, another covariate in the regression model, age and age squared will be collinear, therefore, we centred age before performing the polynomial transformation so that the two would no longer be collinear (46).

#### 3. Genotypes

##### 3.1. Genotyping

Participants were genotyped on the Affymetrix Applied Biosystems UK BiLEVE Axiom Array (N=49,950) and the Affymetrix Applied Biosystems UK Biobank Axiom Array (N=438,427), there is a >95% content overlap between the two chips (47). SNP IDs were annotated using Genome Reference Consortium Human Build 37 (GRCh37/hg19).

##### 3.2. Pre-imputation QC

As described in Bycroft, Freeman (47), SNPs were QC'd for batch effects, plate effects, departures from Hardy-Weinberg equilibrium, sex effects, array effects, and discordance across control replicates. Four of the tests (batch effect, plate effect, departures from HWE, sex effect) were applied to each marker in each batch separately.

##### 3.3. Imputation

Missing genotypes were imputed using the Haplotype Reference Consortium (22) and UK10K haplotype reference (48) panels. As in Legge, Jones (39), SNPs imputed by the UK10K haplotype reference were removed in accordance with guidance from the UK Biobank.

##### 3.4. Post-imputation QC for this study

As previously described in Leonenko, Baker (49), SNPs were removed if (1) they had a MAF less than 1%, (2) they were in deviation from Hardy-Weinberg Equilibrium ($p \leq 10-6$), (3) they were missing for more than 5% of individuals, (4) they had an imputation quality INFO score less then 0.4. Samples were removed if (1) there call rate < 98%, (2) there was evidence of heterozygosity (HET > ±0.1), (3) relatedness based on identity by descent with PI_HAT > 0.2.

We identified a subsample of UKBB participants who had (1) genotype data for rs13107325, (2) had not withdrawn from the study before 25/04/2023, (3) did not have a psychotic spectrum disorder (see above), (4) self-reported White British or Irish ancestry [UKBB Field: 22009], and (5) either attempted the baseline cognitive tasks [UKBB Field: 62] or attempted to complete the Mental Health Questionnaire [UKBB Field: 20400]. Then we excluded one member from each related pair with a kinship coefficient greater than 0.15 and restricted our sample to a genetically homogenous group (see below and Figure 3). We restricted the genetic data to SNPs with INFO score greater then 0.9.

###### 3.4.1. Population stratification

As in Legge, Jones (39), we next used the first five principal components supplied by the UKBB [UKBB Field: 220009] to control population structure by computing a Minimum Covariance Determinant (MCD) estimator of location and scatter. MCD is used to define a hyper-ellipsoid in a multi-dimensional space that contains the majority of MCD points. This is done by finding a subset consisting of 99% of participants for which the covariance matrix had the minimum determinant (MCD points) which represents the volume of highest point density (50, 51). We restricted this hyper-ellipsoid to include MCD points, which represent participants, within the 90$^{th}$ percentile of the MCD distance as this looked to define a group of participants that was genetically homogenous.

#### Statistical Analysis

##### 1. *Missing Data Imputation*

For the Cardiff Schizophrenia Samples, visualisation established that the data could be assumed to be missing at random. Missing phenotype data was imputed, for each sample separately, using multiple imputation by chained equations (MICE) with 100 imputation and ten iterations in the burn-in period. MICE was performed in R using the 'mice' package (52). Performance was assessed by plotting the residuals and convergence of chains; checking the range of imputed values; and comparing the observed and imputed data (53). Each dataset was analysed separately and pooled based on Rubin's rules (54).

##### 2. *Regression Model Assumptions*

As we cannot check the assumptions of the model estimated using the pooled data, we checked the assumptions of five models estimated using the first five imputed datasets. The assumptions of each regression model were checked using the R package performance (55). This package uses a Breusch-Pagan Test to check for heteroscedasticity in linear regression models. Heteroscedasticity in an ordinary least squares model will not bias the regression coefficients, but it will bias the standard error and, consequently, the 95% confidence intervals and significance testing. For phenotypes were there was evidence of statistically significant ($P < .01$) heteroscedasticity according to the Breusch-Pagan Test, we recalculated standard errors, confidence intervals, and P-values using the estimatr::lm_robust function to estimate heteroscedasticity-robust variance, specifically Eicker-Huber-White robust "HC2" (also known as "sandwich") standard errors, as recommended by Astivia and Zumbo (56). No adjustments were made if the assumption of normality was violated. Even in cases of non-normality, beta estimates are unbiased, although P-values could be inaccurate.

### Supplementary Results

#### Cardiff Schizophrenia Samples

##### *1. Descriptive statistics*

In CardiffCOGS, at rs13107325, N=528 participants carried 0 schizophrenia-risk alleles, N=134 carried 1 or 2 schizophrenia-risk alleles (Table 5). No differences were observed between the number of schizophrenia-risk alleles and age at assessment, gender, symptom dimensions, current cognitive ability, years in education, nor educational qualifications. Participants who carried more schizophrenia-risk alleles had lower NART IQ scores (one-way ANOVA F(2) = 3.27, P-value = 0.039), but there was no significant difference in means as estimated by Tukey's HSD test. In Cardiff F-Series, N=344 participants carried 0 schizophrenia-risk alleles, N=78 carried 1 or 2 risk alleles (Table 6). No differences were observed between the number of schizophrenia-risk alleles and any of the phenotypes. In Cardiff SibPairs, N=114 participants carried 0 schizophrenia-risk alleles, N=34 carried 1 or 2 risk alleles (Table 7). No differences were observed between the number of schizophrenia-risk alleles and any of the phenotypes.

##### *2. Missing Data*

72.51% of CardiffCOGS participants, 76.30% of Cardiff F-Series participants, and 58.78% of Cardiff SibPairs participants had complete case data. The proportion of observed (non-missing) data ranged across variables from 69-100% (86-100% CardiffCOGS; 88-100% Cardiff F-Series; 69-100% Cardiff SibPairs; see Table 4). The pattern of missingness is visualised (Figures available upon request). Missing data imputation performed well. Some multicollinearity was identified; PC2 was excluded from imputation models for all three samples, and PC1 from Cardiff F-Series and Cardiff SibPairs. In addition, in the Cardiff SibPairs, education qualification (degree) was excluded when imputing age at interview. For all variables, imputed data were within the range of observed data. Kernel density estimates of the imputed and observed data for linear variables showed comparable distributions (Figures available upon request).

##### *3. Regression models*

###### *3.1. Assumptions*

The assumptions of each regression model were checked using the R package performance (55), see Regression Model Assumptions.

When degree was the dependent variable in the multivariable regression model using the Cardiff SibPairs sample, rs13107325 and a number of PCs had high variance inflation factors (VIF) i.e. VIF ≥ 10. VIF = $(1/1- R^2)$ where $R^2$ is calculated for each independent variable by performing a linear regression of that variable on all the other independent variable. Typically, a high VIF indicates multicollinearity, and the regression betas of these variables may not be accurate. Therefore, we dropped PC5 from the model and as a result the VIF for rs13107325 was consistently < 10. Regression estimates for covariates were not reported to prevent misinterpretation.

###### *3.5. Results stratified by sample*

Univariable (unadjusted) and multivariable (adjusted) regression models using rs13107325 allele count as the independent variable are presented in Table 8 (CardiffCOGS), Table 9 (Cardiff F-Series), and Table 10 (Cardiff Sibpairs). Performance metrics for multivariable regression models for both independent variables are presented in Table 11 (CardiffCOGS), Table 12 (Cardiff F-Series), and Table 13 (Cardiff Sibpairs). $R^2$ and pseudo-$R^2$ were calculated using the R package performance (55) and averaged over the 100 imputed datasets (57).

In the CardiffCOGS sample, rs13107325 was not associated with symptoms dimensions, educational qualifications, or age of psychosis onset, however, possessing more schizophrenia-risk alleles was nominally associated with a lower NART IQ score but did not survive correction for multiple testing (Table 8). In the Cardiff F-Series sample, rs13107325 was not associated with any phenotype (Table 9). In the Cardiff SibPairs sample, carrying more schizophrenia-risk alleles was nominally associated with a higher positive symptom domain score but did not survive correction for multiple testing (Table 10).

#### UK Biobank

##### *1. Descriptive statistics*

At rs13107325, N=304253 participants carried zero schizophrenia-risk alleles, N=48907 carried one, and N=1909 carried two (Table 22). No differences were observed between the number of schizophrenia-risk alleles and age at assessment, years in education, nor psychotic experiences. Male participants were more likely to carry a higher number of schizophrenia-risk alleles than females (Pearson's Chi-squared statistic = 7.40, P-value = 0.030). Participants without GCSEs were more likely to carry a higher number of schizophrenia-risk alleles than those with GCSEs (Pearson's Chi-squared statistic = 20.00, P-value = 0.001). Participants without a degree were more likely to carry a higher number of schizophrenia-risk alleles than those with a degree (Pearson's Chi-squared statistic = 17.00, P-value = 0.001). Participants who carried more schizophrenia-risk alleles had lower g scores (one-way ANOVA F(2) = 7.81, P-value = 0.0004), driven by the difference in means between zero and one risk allele (P-value = 0.003), and lower fluid intelligence scores (one-way ANOVA F(2) = 24.10, P-value = <0.001), driven by the differences in means between all three allele-carrier groups (0 v 1, P-value = <0.001; 0 v 2, P-value = <0.001; 1 v 2, P-value = 0.001). These differences, while statistically significant were very small, e.g. 53.84% of participants with 0 risk alleles were female, 53.19% with 1 risk alleles, and 53.27% with 2 risk alleles.

##### *2. Regression Models*

###### *2.1. Assumptions*

The assumptions of each regression model were checked using the R package performance (55), see Regression Model Assumptions.

When g score and fluid intelligence were used as the dependent variables in the multivariable regression models, both age at assessment and sex had high variance inflation factors (VIF) i.e. VIF ≥ 10. VIF = (1/1- $R^2$) where $R^2$ is calculated for each independent variable by performing a linear regression of that variable on all the other independent variable. Typically, a high VIF indicates multicollinearity, and the regression betas of these variables may not be accurate. However, because age at assessment and sex variables were included to account for confounding, we did not take any further steps to mitigate the presence of multicollinearity. Regression estimates for covariates were not reported to prevent misinterpretation.
