## Supplementary Material Tables for "SLC39A8.p.(Ala391Thr) is associated with poorer cognitive ability: a cross-sectional study of schizophrenia and the general UK population"

### Supplementary Tables

#### Table of Tables

Table 1. rs13107325 in GWAS Summary Statistics.

| GWAS | PMID | CHR | SNP | BP | A1 | A2 | MEASURE OF EFFECT | EFFECT SIZE | SE | P | NCAS | NCON | N |
|---|---|---|---|---|---|---|---|---|---|---|---|---|---|
| ADHD 2022 | 36702997[1] | 4 | rs13107325 | 103188709 | C | T | OR | 1.00 | 0.02 | 8.44E-01 | 38691 | 186843 | NA |
| Alcohol dependence 2018 | 30482948[1] | 4 | rs13107325 | 103188709 | T | C | BETA | -0.02 | 0.05 | 6.20E-01 | | | |
| Alcohol use/AUDIT 2019 | 30336701[1] | | SNP NOT FOUND | | | | | | | | | | |
| Alzheimer's Disease 2021 | 34493870[1] | 4 | | 103188709 | T | C | Z | 1.33 | NA | 1.83E-01 | | | 726258 |
| Anxiety Disorders 2016 | 26754954[1] | 4 | rs13107325 | 103188709 | T | C | EFFECT | 0.07 | 0.07 | 3.66E-01 | | | 7440 |
| Autism Spectrum Disorder 2019 | 30804558[1] | 4 | rs13107325 | 103188709 | T | C | OR | 1.05 | 0.03 | 1.09E-01 | | | |
| Bipolar 2021 | 34002096[1] | 4 | rs13107325 | 103188709 | C | T | BETA | -0.05 | 0.02 | 1.21E-02 | 41816 | 371234 | |
| Cannabis use disorder 2020 | 33096046[1] | 4 | rs13107325 | 103188709 | T | C | Z | 0.83 | NA | 4.08E-01 | 16153 | 350362 | 366515 |
| Eating Disorders 2019 (A1=REF, A2=ALT) | 31308545[1] | 4 | rs13107325 | 103188709 | T | C | BETA | 0.00 | 0.03 | 9.38E-01 | | | |
| Educational Attainment 2022 | 35361970[2] | 4 | rs13107325 | 103188709 | T | C | BETA | -0.02 | 0.00 | 1.08E-13 | | | |
| Intelligence 2018 | 29942086[3] | 4 | rs13107325 | 103188709 | T | C | Z | -9.49 | 0.01 | 2.23E-21 | | | 265961 |
| Major Depressive Disorder 2018 | 29700475[1] | 4 | rs13107325 | 103188709 | T | C | OR | 0.97 | 0.02 | 1.62E-01 | 45396 | 97250 | |
| Major Depressive Disorder 2019 only 10000 SNPs | 30718901[1] | | SNP NOT FOUND | | | | | | | | | | |
| Major Depressive Disorder 2021 with UKB | 34586374[1] | | SNP NOT FOUND | | | | | | | | | | |
| Major Depressive Disorder 2021 without UKB | 34586374[1] | | SNP NOT FOUND | | | | | | | | | | |
| OCD 2018 | 28761083[1] | 4 | rs13107325 | 103188709 | T | C | OR | 1.06 | 0.06 | 3.50E-01 | | | |
| Opioid dependence 2020 opioid-exposed_vs._opioid-unexposed | 32099098[1] | | rs13107325 | | T | C | Z | 0.28 | | 7.80E-01 | 3515 | 14404 | 17919 |
| Opioid dependence 2020 OD_cases_vs._opioid-unexposed | 32099098[1] | | rs13107325 | | T | C | Z | -0.46 | | 6.44E-01 | 3520 | 13320 | 16840 |
| Panic Disorder 2019 | 31712720[1] | 4 | rs13107325 | 103188709 | T | C | BETA | 0.12 | 0.08 | 1.47E-01 | 2147 | 7760 | |
| Parkinson's Disease 2019 | 31701892[4] | 4 | | 103188709 | T | C | BETA | -0.03 | 0.03 | 3.31E-01 | 33674 | 449056 | |
| Post Traumatic Stress Disorder 2019 | 31594949[1] | 4 | rs13107325 | 103188709 | T | C | OR | 1.11 | 0.03 | 7.75E-05 | 28818 | 165527 | |

| | | | | | | | | | | | | |
|---|---|---|---|---|---|---|---|---|---|---|---|---|
| Schizophrenia 2022 (A1=REF, A2=ALT) | 35396580[1] | 4 | rs13107325 | 103188709 | C | T | BETA | -0.16 | 0.02 | 1.92E-21 | 74776 | 101023 |
| Tourette Syndrome 2019 | 30818990[1] | 4 | rs13107325 | 103188709 | T | C | OR | 0.99 | 0.05 | 9.04E-01 | | |

Note: (a) P-values shown in bold indicate an association that is significant at the genome-wide significance threshold. (b) On Parkinson's Disease, Pickrell, Berisa (1) reported a nominally significant association with rs13107325 but this was based on data from 23andMe and is not replicated in the GWAS of cases ascertained by clinicians reported by Nalls, Blauwendraat (2) and included in the Table. (c) Genome-wide significant p-values are in bold.

[1] https://pgc.unc.edu/for-researchers/download-results/

[2] https://thessgac.com/papers/

[3] https://ctg.cncr.nl/software/summary_statistics/

[4] https://drive.google.com/drive/folders/10bGj6HfAXgl-JslpI9ZJIL_JIgZyktxn

Cardiff Schizophrenia Samples

#### 1. *Ancestry*

Table 2. The proportion of participants in the Cardiff Schizophrenia Samples, within each biogeographical ancestry, stratified by rs13107325 allele count and sample.

| | | **Biogeographical ancestry groups** | | | | | | | | | | | | | | | |
|---|---|---|---|---|---|---|---|---|---|---|---|---|---|---|---|---|---|
| **Study** | **rs13107325 schizophrenia-risk allele count** | **African American / Afro-Caribbean** | | **Central / South Asian** | | **East Asian** | | **European** | | **Middle Eastern / North African** | | **Mixed** | | **Sub-Saharan African** | | **Total** | |
| CardiffCOGS | 0 | 1 | 100.00% | 4 | 80.00% | 1 | 100.00% | 522 | 79.69% | 1 | 100.00% | 1 | 100.00% | 1 | 100.00% | 531 | 79.85% |
| | 1 | 0 | 0.00% | 1 | 20.00% | 0 | 0.00% | 128 | 19.54% | 0 | 0.00% | 0 | 0.00% | 0 | 0.00% | 129 | 19.40% |
| | 2 | 0 | 0.00% | 0 | 0.00% | 0 | 0.00% | 5 | 0.76% | 0 | 0.00% | 0 | 0.00% | 0 | 0.00% | 5 | 0.75% |
| | Total | 1 | | 5 | | 1 | | 655 | | 1 | | 1 | | 1 | | 665 | |
| F-Series | 0 | 3 | 75.00% | 5 | 100.00% | 0 | - | 335 | 81.51% | 1 | 50.00% | 2 | 100.00% | 2 | 100.00% | 348 | 81.69% |
| | 1 | 1 | 25.00% | 0 | 0.00% | 0 | - | 72 | 17.52% | 1 | 50.00% | 0 | 0.00% | 0 | 0.00% | 74 | 17.37% |
| | 2 | 0 | 0.00% | 0 | 0.00% | 0 | - | 4 | 0.97% | 0 | 0.00% | 0 | 0.00% | 0 | 0.00% | 4 | 0.94% |
| | Total | 4 | | 5 | | 0 | | 411 | | 2 | | 2 | | 2 | | 426 | |
| SibPairs | 0 | 0 | 0.00% | 3 | 100.00% | 0 | - | 111 | 77.08% | 0 | - | 1 | 100.00% | 2 | 100.00% | 117 | 77.48% |
| | 1 | 1 | 100.00% | 0 | 0.00% | 0 | - | 30 | 20.83% | 0 | - | 0 | 0.00% | 0 | 0.00% | 31 | 20.53% |
| | 2 | 0 | 0.00% | 0 | 0.00% | 0 | - | 3 | 2.08% | 0 | - | 0 | 0.00% | 0 | 0.00% | 3 | 1.99% |
| | Total | 1 | | 3 | | 0 | | 144 | | 0 | | 1 | | 2 | | 151 | |

Note: (a) Because biogeographical ancestry and rs13107325 allele count cannot be easily obtained there is assumed to be no meaningful disclosure risk. (b) No participants were classified as American, Oceanian, Latino. (c) The mixed group consists of participants who did not pass thresholds for any one of the pre-defined ancestries. (d) The Middle Eastern/North African ancestry is referred to as Near Eastern in Huddart, Fohner (3). (e) Within each study we performed two-sided Fisher's Exact tests, to test whether the frequency of allele counts differed across ancestry groups: CardiffCOGS P = 1, F-Series P = 0.6, SibPairs = 0.5. (f) Within each study we performed two-sided Fisher's Exact tests, after transforming the allele count into a binary variable (no schizophrenia alelles vs at least one schizophrenia allele): CardiffCOGS P = 1, F-Series P = 0.5, SibPairs = 0.5.

Table 3. Kruskal-Wallis rank sum tests comparing rs13107325 allele count against biogeographical ancestry probabilities in the Cardiff Schizophrenia Samples.

| **Study** | | | **CardiffCOGS (N=665)** | | | **F-Series (N=426)** | | | **SibPairs (N=151)** | | |
|---|---|---|---|---|---|---|---|---|---|---|---|
| **rs13107325 schizophrenia-allele count** | | | **0** | **1** | **2** | **0** | **1** | **2** | **0** | **1** | **2** |
| Biogeographical ancestry probabilities | African American/Afro-Caribbean | Mean (sd) | 0.003 (0.046) | 0 (0) | 0 (0) | 0.007 (0.081) | 0.014 (0.116) | 0 (0) | 0.001 (0.008) | 0.031 (0.170) | 0 (0) |
| | | Chi-squared statistic | 0.5 | | | 2 | | | 4.1 | | |
| | | P-Value | 0.8 | | | 0.7 | | | 0.4 | | |
| | East Asian | Mean (sd) | 0.001 (0.032) | 0 (0) | 0 (0) | 0.00003 (0.001) | 0 (0) | 0 (0) | 0.002 (0.019) | 0 (0) | 0 (0) |
| | | Chi-squared statistic | 0.5 | | | 0.45 | | | 0.29 | | |
| | | P-Value | 0.8 | | | 0.8 | | | 0.6 | | |
| | European | Mean (sd) | 0.985 (0.113) | 0.992 (0.088) | 1 (0) | 0.965 (0.175) | 0.973 (0.161) | 1 (0) | 0.951 (0.207) | 0.968 (0.179) | 0 (0) |
| | | Chi-squared statistic | 2.1 | | | 7.6 | | | 5.9 | | |
| | | P-Value | 1 | | | 0.9 | | | 0.4 | | |
| | Near Eastern | Mean (sd) | 0.002 (0.030) | 0 (0) | 0 (0) | 0.005 (0.058) | 0.013 (0.114) | 0 (0) | 0.004 (0.041) | 0.002 (0.010) | 0 (0) |
| | | Chi-squared statistic | 1 | | | 6.6 | | | 3.8 | | |
| | | P-Value | 0.9 | | | 0.7 | | | 0.3 | | |
| | Central/South Asian | Mean (sd) | 0.007 (0.083) | 0.008 (0.088) | 0 (0) | 0.015 (0.117) | 0 (0) | 0 (0) | 0.026 (0.159) | 0 (0) | 0 (0) |
| | | Chi-squared statistic | 0.81 | | | 1.4 | | | 1.2 | | |
| | | P-Value | 0.9 | | | 0.7 | | | 0.6 | | |
| | Sub-Saharan African | Mean (sd) | 0.002 (0.043) | 0 (0) | 0 (0) | 0.007 (0.079) | 0 (0) | 0 (0) | 0.016 (0.125) | 0 (0) | 0 (0) |
| | | Chi-squared statistic | 0.25 | | | 0.67 | | | 0.58 | | |
| | | P-Value | 0.6 | | | 0.9 | | | 0.7 | | |

Note: (a) Because biogeographical ancestry and rs13107325 allele count cannot be easily obtained there is assumed to be no meaningful disclosure risk. (b) The Kruskal-Wallis rank sum test is a non-parametric alternative to the one-way ANOVA test. No participants were classified as American, Oceanian, Latino.
Abbreviations: sd, standard deviation.

#### 2. *Missing data*

Table 4. Proportion of non-missing data in the Cardiff Schizophrenia Samples.

| | CardiffCOGS | | F-Series | | SibPairs | |
|---|---|---|---|---|---|---|
| | **N** | **%** | **N** | **%** | **N** | **%** |
| N | 662 | | 442 | | 148 | |
| Age at interview | 662 | 100.00 | 408 | 92.31 | 105 | 70.95 |
| Gender | 662 | 100.00 | 413 | 93.44 | 148 | 100.00 |
| Three-factor symptom dimensions | | | | | | |
| Positive symptoms | 639 | 96.53 | 396 | 89.59 | 139 | 93.92 |
| Negative symptoms of diminished expressivity | 639 | 96.53 | 396 | 89.59 | 139 | 93.92 |
| Disorganised symptoms | 639 | 96.53 | 396 | 89.59 | 139 | 93.92 |
| Five-factor symptom dimensions | | | | | | |
| Positive symptoms | 604 | 91.24 | NA | NA | NA | NA |
| Negative symptoms of diminished expressivity | 604 | 91.24 | NA | NA | NA | NA |
| Disorganised symptoms | 604 | 91.24 | NA | NA | NA | NA |
| Negative symptoms of motivation and pleasure | 604 | 91.24 | NA | NA | NA | NA |
| Cognitive ability | 604 | 91.24 | NA | NA | NA | NA |
| NART IQ | 575 | 86.86 | NA | NA | NA | NA |
| Years in education | 642 | 96.98 | 401 | 90.72 | 103 | 69.59 |
| Educational qualification: GCSEs | 649 | 98.04 | 392 | 88.69 | 103 | 69.59 |
| Educational qualification: degree | 649 | 98.04 | 392 | 88.69 | 103 | 69.59 |
| Age at psychosis onset | 637 | 96.22 | 397 | 89.82 | 135 | 91.22 |
| Principle components | 662 | 100.00 | 442 | 100.00 | 148 | 100.00 |

#### 3. Descriptive statistics

Table 5. Descriptive statistics using the observed data from the CardiffCOGS sample.

| Phenotypes | rs13107325 schizophrenia-risk allele count | | | | |
|---|---|---|---|---|---|
| | 0 | 1 | 2 | Total | P-Value[1] |
| Age at interview | | | | | |
| Mean (sd) | 43.34 (11.94) | 43.23 (12.76) | 42.20 (15.37) | 43.31 (12.11) | 0.976 |
| N Missing | 0 | 0 | 0 | 0 | |
| Gender | | | | | |
| Male | 347 (65.72%) | 85 (65.89%) | - | 432 (65.75%) | 0.482 |
| Female | 181 (34.28%) | 44 (34.11%) | - | 225 (34.25%) | |
| N Missing | 0 | 0 | - | 0 | |
| Three-factor symptom dimensions | | | | | |
| Positive symptoms | | | | | |
| Mean (sd) | 0.05 (0.34) | -0.02 (0.28) | 0.15 (0.42) | 0.04 (0.33) | 0.127 |
| N Missing | 15 | 8 | 0 | 23 | |
| Negative symptoms of diminished expressivity | | | | | |
| Mean (sd) | 0 (0.51) | 0.04 (0.52) | -0.2 (0.4) | 0 (0.51) | 0.499 |
| N Missing | 15 | 8 | 0 | 23 | |
| Disorganised symptoms | | | | | |
| Mean (sd) | 0.04 (0.26) | 0.04 (0.27) | -0.06 (0.12) | 0.04 (0.26) | 0.691 |
| N Missing | 15 | 8 | 0 | 23 | |
| Five-factor symptom dimensions | | | | | |
| Positive symptoms | | | | | |
| Mean (sd) | 0.06 (0.49) | 0 (0.4) | 0.25 (0.63) | 0.05 (0.48) | 0.288 |
| N Missing | 45 | 13 | 0 | 58 | |
| Negative symptoms of diminished expressivity | | | | | |
| Mean (sd) | -0.04 (0.49) | -0.03 (0.51) | 0.01 (0.54) | -0.04 (0.49) | 0.973 |
| N Missing | 45 | 13 | 0 | 58 | |

| | | | | | |
|---|---|---|---|---|---|
| Disorganised symptoms | | | | | |
| Mean (sd) | 0.01 (0.30) | 0 (0.30) | -0.1 (0.18) | 0.01 (0.3) | 0.624 |
| N Missing | 45 | 13 | 0 | 58 | |
| Negative symptoms of motivation and pleasure | | | | | |
| Mean (sd) | -0.05 (0.60) | -0.01 (0.62) | -0.22 (0.54) | -0.04 (0.6) | 0.669 |
| N Missing | 45 | 13 | 0 | 58 | |
| Cognitive ability | | | | | |
| Mean (sd) | 0.02 (0.82) | -0.13 (0.82) | -0.19 (0.48) | -0.01 (0.82) | 0.211 |
| N Missing | 45 | 13 | 0 | 58 | |
| NART IQ | | | | | |
| Mean (sd) | 0.05 (0.97) | -0.15 (1.08) | -0.75 (0.91) | 0 (1.00) | 0.039 |
| N Missing | 77 | 10 | 0 | 87 | |
| Years in education | | | | | |
| Mean (sd) | 12.77 (2.56) | 12.64 (2.69) | 11.4 (1.52) | 12.74 (2.58) | 0.444 |
| N Missing | 17 | 3 | 0 | 20 | |
| Educational qualification | | | | | |
| No GCSE | 185 (35.71%) | 52 (41.27%) | - | 237 (36.80%) | 0.505 |
| GCSE | 333 (64.29%) | 74 (58.73%) | - | 407 (63.20%) | |
| N Missing | 10 | 3 | - | 13 | |
| Educational qualification | | | | | |
| No Degree | 453 (87.45%) | 106 (84.13%) | - | 559 (86.80%) | 0.418 |
| Degree | 65 (12.55%) | 20 (15.87%) | - | 85 (13.20%) | |
| N Missing | 10 | 3 | - | 13 | |
| Age at psychosis onset | | | | | |
| Mean (sd) | 24.57 (8.78) | 24.45 (9.62) | 28.2 (12.68) | 24.57 (8.97) | 0.657 |
| N Missing | 21 | 4 | 0 | 25 | |

Note: (a) Because there is a meaningful disclosure risk, non-missing counts <5 have been supressed and totals recalculated. (b) [1] Analysis of variance (ANOVA) were used to test continuous variables and Pearson's Chi-squared tests for categorical variables.
Abbreviations: sd, standard deviation.

Table 6. Descriptive statistics using the observed data from the Cardiff F-Series sample.

| Phenotypes | rs13107325 schizophrenia-risk allele count | | | | |
|---|---|---|---|---|---|
| | 0 | 1 | 2 | Total | P-Value[1] |
| Age at interview | | | | | |
| Mean (sd) | 41.90 (14.18) | 43.61 (13.59) | 33.75 (13.07) | 42.11 (14.08) | 0.319 |
| N Missing | 10 | 4 | 0 | 14 | |
| Gender | | | | | |
| Male | 234 (69.64%) | 55 (75.34%) | - | 289 (70.66%) | 0.417 |
| Female | 102 (30.36%) | 18 (24.66%) | - | 120 (29.34%) | |
| N Missing | 8 | 1 | - | 9 | |
| Three-factor symptom dimensions | | | | | |
| Positive symptoms | | | | | |
| Mean (sd) | 0.01 (0.12) | 0 (0.13) | 0.03 (0.18) | 0.01 (0.12) | 0.852 |
| N Missing | 21 | 4 | 1 | 26 | |
| Negative symptoms of diminished expressivity | | | | | |
| Mean (sd) | 0.02 (0.53) | 0.02 (0.55) | 0.20 (0.31) | 0.02 (0.53) | 0.846 |
| N Missing | 21 | 4 | 1 | 26 | |
| Disorganised symptoms | | | | | |
| Mean (sd) | -0.01 (0.31) | 0.02 (0.33) | 0.06 (0.19) | 0 (0.31) | 0.737 |
| N Missing | 21 | 4 | 1 | 26 | |
| Years in education | | | | | |
| Mean (sd) | 12.32 (1.55) | 11.88 (2.03) | 12.67 (1.15) | 12.24 (1.64) | 0.128 |
| N Missing | 14 | 6 | 1 | 21 | |
| Educational qualification | | | | | |
| No GCSE | 131 (40.56%) | 31 (47.69%) | - | 162 (41.75%) | 0.451 |
| GCSE | 192 (59.44%) | 34 (52.31%) | - | 226 (58.25%) | |
| N Missing | 21 | 9 | - | 30 | |
| Educational qualification | | | | | 0.527 |
| No Degree | 281 (87.00%) | 59 (90.77%) | - | 340 (87.63%) | |

| | | | | | |
|---|---|---|---|---|---|
| Degree | 42 (13.00%) | 6 (9.23%) | - | 48 (12.37%) | |
| N Missing | 21 | 9 | - | 30 | |
| Age at psychosis onset | | | | | |
| Mean (sd) | 24.37 (8.98) | 24.44 (9.34) | 18.50 (2.89) | 24.32 (9.01) | 0.431 |
| N Missing | 21 | 4 | 0 | 25 | |

Note: (a) Because there is a meaningful disclosure risk, non-missing counts <5 have been supressed and totals recalculated. (b) [1] Analysis of variance (ANOVA) were used to test continuous variables and Pearson's Chi-squared tests for categorical variables.
Abbreviations: sd, standard deviation.

Table 7. Descriptive statistics using the observed data from the Cardiff SibPairs sample.

| Phenotypes | rs13107325 schizophrenia-risk allele count | | | | |
|---|---|---|---|---|---|
| | 0 | 1 | 2 | Total | P-Value[1] |
| Age at interview | | | | | |
| Mean (sd) | 42.23 (12.96) | 39.77 (13.52) | 40.00 (1.41) | 41.68 (12.93) | 0.722 |
| N Missing | 33 | 9 | 1 | 43 | |
| Gender | | | | | |
| Male | 76 (66.67%) | 20 (64.52%) | - | 96 (66.21%) | 0.457 |
| Female | 38 (33.33%) | 11 (35.48%) | - | 49 (33.79%) | |
| N Missing | 0 | 0 | - | 0 | |
| Three-factor symptom dimensions | | | | | |
| Positive symptoms | | | | | |
| Mean (sd) | -0.04 (0.26) | 0.08 (0.31) | 0.16 (0.30) | -0.01 (0.27) | 0.072 |
| N Missing | 5 | 4 | 0 | 9 | |
| Negative symptoms of diminished expressivity | | | | | |
| Mean (sd) | 0.07 (0.56) | 0.05 (0.59) | 0.19 (0.68) | 0.07 (0.56) | 0.925 |
| N Missing | 5 | 4 | 0 | 9 | |
| Disorganised symptoms | | | | | |
| Mean (sd) | 0.01 (0.35) | 0.11 (0.46) | 0.20 (0.42) | 0.03 (0.38) | 0.334 |
| N Missing | 5 | 4 | 0 | 9 | |
| Years in education | | | | | |
| Mean (sd) | 11.42 (1.06) | 11.55 (0.74) | 11.50 (0.71) | 11.45 (0.99) | 0.866 |
| N Missing | 35 | 9 | 1 | 45 | |
| Educational qualification | | | | | |
| No GCSE | 48 (61.54%) | 14 (63.64%) | - | 62 (62.00%) | 0.971 |
| GCSE | 30 (38.46%) | 8 (36.36%) | - | 38 (38.00%) | |
| N Missing | 36 | 9 | 0 | 45 | |
| Educational qualification | | | | | |
| No Degree | 77 (98.72%) | 22 (100.00%) | - | 99 (99.00%) | 0.851 |

| | | | | | |
|---|---|---|---|---|---|
| Degree | - | - | - | 1 (1.00%) | |
| N Missing | 36 | 9 | 0 | 45 | |
| Age at psychosis onset | | | | | |
| Mean (sd) | 24.10 (7.77) | 22.68 (5.42) | 23.00 (13.89) | 23.78 (7.46) | 0.663 |
| N Missing | 10 | 3 | 0 | 13 | |

Note: (a) Because there is a meaningful disclosure risk, non-missing counts <5 have been supressed and totals recalculated. (b) [1] Analysis of variance (ANOVA) were used to test continuous variables and Pearson's Chi-squared tests for categorical variables.
Abbreviations: sd, standard deviation.

#### *4. Regression analyses*

Table 8. Regression analyses using the observed/imputed data from the CardiffCOGS sample (N=662) and rs13107325 allele count as the independent variable.

| | **Univariable** | | | | | **Multivariable[1]** | | | | | |
|---|---|---|---|---|---|---|---|---|---|---|---|
| | **Beta/OR** | **Lower 95% CI** | **Upper 95% CI** | **Standard Error** | **P-Value** | **Beta/OR** | **Lower 95% CI** | **Upper 95% CI** | **Standard Error** | **P-Value** | **P-Value adj.** |
| Three-factor symptom dimensions | | | | | | | | | | | |
| Positive symptoms | *-0.05* | *-0.11* | *0.02* | 0.03 | 0.144 | *-0.05* | *-0.11* | *0.01* | 0.03 | 0.130 | 0.561 |
| Negative symptoms of diminished expressivity | *0.02* | *-0.08* | *0.11* | 0.05 | 0.714 | *0.02* | *-0.08* | *0.11* | 0.05 | 0.728 | 0.789 |
| Disorganised symptoms | *-0.01* | *-0.06* | *0.04* | 0.02 | 0.705 | *-0.01* | *-0.06* | *0.04* | 0.02 | 0.682 | 0.887 |
| Five-factor symptom dimensions | | | | | | | | | | | |
| Positive symptoms | *-0.05* | *-0.13* | *0.04* | 0.04 | 0.301 | *-0.05* | *-0.14* | *0.04* | 0.04 | 0.264 | 0.686 |
| Negative symptoms of diminished expressivity | *0.00* | *-0.09* | *0.09* | 0.05 | 0.972 | *0.00* | *-0.09* | *0.09* | 0.05 | 0.991 | 0.991 |
| Disorganised symptoms | *0.02* | *-0.09* | *0.13* | 0.06 | 0.697 | *-0.02* | *-0.07* | *0.04* | 0.03 | 0.520 | 0.967 |
| Negative symptoms of motivation and pleasure | *-0.02* | *-0.07* | *0.04* | 0.03 | 0.543 | *0.02* | *-0.09* | *0.13* | 0.05 | 0.711 | 0.840 |
| Cognitive ability | *-0.12* | *-0.27* | *0.03* | 0.08 | 0.117 | *-0.12* | *-0.26* | *0.02* | 0.07 | 0.095 | 0.616 |
| NART IQ | *-0.22* | *-0.40* | *-0.03* | 0.09 | 0.020 | *-0.21* | *-0.39* | *-0.03* | 0.09 | 0.021 | 0.274 |
| Years in education | *-0.20* | *-0.66* | *0.27* | 0.24 | 0.410 | *-0.16* | *-0.61* | *0.30* | 0.23 | 0.500 | 1.082 |
| Educational qualification: GCSE | 0.80 | 0.56 | 1.16 | 0.19 | 0.239 | 0.78 | 0.54 | 1.14 | 0.19 | 0.201 | 0.653 |
| Educational qualification: degree | 1.16 | 0.69 | 1.94 | 0.26 | 0.574 | 1.18 | 0.70 | 1.98 | 0.27 | 0.536 | 0.871 |
| Age at psychosis onset | *0.39* | *-1.27* | *2.04* | 0.84 | 0.647 | *0.45* | *-1.01* | *1.91* | 0.74 | 0.544 | |
| Age at psychosis onset[2] | | | | | | *0.45* | *-1.05* | *1.95* | 0.763 | 0.554 | 0.800 |

Note: (a) Ordinary least square regression models. (b) Beta coefficients indicated by italic. (c) [1] adjusted for age at first interview, gender, and principal components (PC)1-5. (d) [2] Recalculated standard errors, confidence intervals, and p-values to estimate heteroscedasticity-robust variance. (e) No False Discovery Rate (FDR) adjusted p-value (adjusted for 13 tests) was statistically significant at an alpha level ≤ .05.
Abbreviations: CI, confidence interval; OR, odds ratio.

Table 9. Regression analyses using the observed/imputed data from the Cardiff F-Series sample (N=422) and rs13107325 allele count as the independent variable.

| | **Univariable** | | | | | **Multivariable[1]** | | | | | |
|---|---|---|---|---|---|---|---|---|---|---|---|
| | **Beta/OR** | **Lower 95% CI** | **Upper 95% CI** | **Standard Error** | **P-Value** | **Beta/OR** | **Lower 95% CI** | **Upper 95% CI** | **Standard Error** | **P-Value** | **P-Value adj.** |
| Three-factor symptom dimensions | | | | | | | | | | | |
| Positive symptoms | *-0.01* | *-0.03* | *0.02* | 0.02 | 0.734 | *0.00* | *-0.03* | *0.03* | 0.02 | 0.857 | 0.857 |
| Negative symptoms of diminished expressivity | *0.02* | *-0.11* | *0.14* | 0.06 | 0.765 | *0.03* | *-0.10* | *0.15* | 0.07 | 0.698 | 0.977 |
| Disorganised symptoms | *0.03* | *-0.04* | *0.10* | 0.04 | 0.431 | *0.03* | *-0.05* | *0.11* | 0.04 | 0.423 | 1.482 |
| Years in education | *-0.34* | *-0.73* | *0.06* | 0.20 | 0.098 | *-0.29* | *-0.69* | *0.10* | 0.20 | 0.140 | |
| Years in education[2] | | | | | | *-0.29* | *-0.73* | *0.14* | 0.22 | 0.185 | 1.292 |
| Educational qualification: GCSE | 0.85 | 0.53 | 1.38 | 0.24 | 0.512 | 0.89 | 0.53 | 1.50 | 0.27 | 0.661 | 1.157 |
| Educational qualification: degree | 0.92 | 0.43 | 1.94 | 0.38 | 0.817 | 0.81 | 0.37 | 1.77 | 0.40 | 0.598 | 1.395 |
| Age at psychosis onset | *-0.53* | *-2.62* | *1.56* | 1.06 | 0.615 | *-0.30* | *-2.14* | *1.54* | 0.94 | 0.752 | |
| Age at psychosis onset[2] | | | | | | *-0.30* | *-2.14* | *1.55* | 0.94 | 0.752 | 0.877 |

Note: (a) Ordinary least square regression models. (b) Beta coefficients indicated by italic. (c) [1] Adjusted for age at first interview, gender, and principal components (PC)1-5, 6, and 9. (d) [2] Recalculated standard errors, confidence intervals, and p-values to estimate heteroscedasticity-robust variance. (e) No False Discovery Rate (FDR) adjusted p-value (adjusted for 7 tests) was statistically significant at an alpha level ≤ .05.
Abbreviations: CI, confidence interval; OR, odds ratio.

Table 10. Regression analyses using the observed/imputed data from the Cardiff SibPairs sample (N=148) and rs13107325 allele count as the independent variable.

| | **Univariable** | | | | | **Multivariable[1]** | | | | | |
|---|---|---|---|---|---|---|---|---|---|---|---|
| | **Beta/OR** | **Lower 95% CI** | **Upper 95% CI** | **Standard Error** | **P-Value** | **Beta/OR** | **Lower 95% CI** | **Upper 95% CI** | **Standard Error** | **P-Value** | **P-Value adj.** |
| Three-factor symptom dimensions | | | | | | | | | | | |
| Positive symptoms | *0.11* | *0.02* | *0.21* | 0.05 | 0.021 | *0.11* | *0.01* | *0.20* | 0.05 | 0.025 | 0.172 |
| Negative symptoms of diminished expressivity | *0.00* | *-0.20* | *0.20* | 0.10 | 0.987 | *0.01* | *-0.19* | *0.21* | 0.10 | 0.907 | 1.058 |
| Disorganised symptoms | *0.10* | *-0.03* | *0.24* | 0.07 | 0.140 | *0.12* | *-0.02* | *0.25* | 0.07 | 0.093 | 0.327 |
| Years in education | *0.16* | *-0.22* | *0.55* | 0.20 | 0.402 | *0.13* | *-0.24* | *0.49* | 0.18 | 0.495 | 0.867 |
| Educational qualification: GCSE | -0.17 | -0.96 | 0.61 | 0.40 | 0.666 | 0.78 | 0.34 | 1.78 | 0.41 | 0.557 | 0.779 |
| Educational qualification: degree | -1.90 | -1110.31 | 1106.51 | 560.77 | 0.997 | 0.07 | 0.00 | Inf | 665.02 | 0.997 | 0.997 |
| Age at psychosis onset | *-1.10* | *-3.72* | *1.53* | 1.33 | 0.410 | *-0.99* | *-3.50* | *1.53* | 1.27 | 0.438 | |
| Age at psychosis onset[2] | | | | | | *-0.99* | *-3.49* | *1.51* | 1.26 | 0.435 | 1.015 |

Note: (a) Ordinary least square regression models. (b) Beta coefficients indicated by italic. (c) For the degree-dependent variable model, the predicted probabilities of one or more observations in the data were indistinguishable from 0 or 1 possibly due to the small sample size. (d) [1] Adjusted for age at first interview, gender, and principal components (PC)1-5, except for the degree-dependent variable model where PC3 and PC5 was removed due to multicollinearity. (e) [2] Recalculated standard errors, confidence intervals, and p-values to estimate heteroscedasticity-robust variance. (f) No False Discovery Rate (FDR) adjusted p-value (adjusted for 7 tests) was statistically significant at an alpha level ≤ .05. Abbreviations: CI, confidence interval; OR, odds ratio.

Table 11. Performance metrics for regression models using the observed/imputed data from the CardiffCOGS sample (N=662) and rs13107325 allele count as the independent variables.

| | rs13107325[1] | | | |
|---|---|---|---|---|
| | **Tjur's $R^2$** | **$R^2$ (SNP)** | **$R^2$ adj. (SNP)** | **RMSE** |
| Three-factor symptom dimensions | | | | |
| Positive symptoms | | 0.89% (0.33%) | -0.32% (0.18%) | 0.3302 |
| Negative symptoms of diminished expressivity | | 2.36% (0.01%) | 1.17% (-0.14%) | 0.5015 |
| Disorganised symptoms | | 0.74% (0.01%) | -0.48% (-0.14%) | 0.2570 |
| Five-factor symptom dimensions | | | | |
| Positive symptoms | | 1.05% (0.20%) | -0.16% (0.05%) | 0.4690 |
| Negative symptoms of diminished expressivity | | 3.05% (0.00%) | 1.86% (-0.15%) | 0.4901 |
| Disorganised symptoms | | 0.64% (0.03%) | -0.58% (-0.12%) | 0.2972 |
| Negative symptoms of motivation and pleasure | | 2.98% (0.01%) | 1.79% (-0.14%) | 0.5811 |
| Cognitive ability | | 17.19% (0.11%) | 16.18% (-0.02%) | 0.7429 |
| NART IQ | | 7.51% (1.12%) | 6.38% (0.99%) | 0.9593 |
| Years in education | | 5.82% (0.06%) | 4.66% (-0.08%) | 2.4914 |
| Educational qualification: GCSE | 4.12% (0.20%) | | | 0.4728 |
| Educational qualification: degree | 1.33% (0.04%) | | | 0.3354 |
| Age at psychosis onset | | 24.37% (0.08%) | 23.44% (-0.04%) | 7.9080 |

Note: (a) Tjur's $R^2$ is an alternative to other pseudo-$R^2$ values like Nagelkerke's $R^2$ or Cox-Snell $R^2$ and can be read like any other (pseudo-)$R^2$ value (4). (b) RMSE is the square root of the variance of the residuals and indicates the absolute fit of the model to the data (difference between observed data to model's predicted values). It can be interpreted as the standard deviation of the unexplained variance, and is in the same units as the response variable. Lower values indicate better model fit. (c) (SNP) is the difference between the variance explained by the full model and the variance explained by a covariate-only model and can thus be interpreted as the variance explained by the SNP alone. (d) [1] Model adjusted for age at first interview, gender, and principal components (PC)1-5. The difference between the variance of the model containing rs13107325 adjusted for age at first interview, gender, and principal components (PC)1-5 and the model containing only the covariates is shown in brackets.

Table 12. Performance metrics for regression models using the observed/imputed data from the Cardiff F-Series sample (N=422) and rs13107325 allele count as the independent variables.

| | rs13107325[1] | | | |
|---|---|---|---|---|
| | **Tjur's $R^2$ (SNP)** | **$R^2$ (SNP)** | **$R^2$ adj. (SNP)** | **RMSE** |
| Three-factor symptom dimensions | | | | |
| Positive symptoms | | 3.95% (0.73%) | 1.62% (0.03%) | 0.1212 |
| Negative symptoms of diminished expressivity | | 0.92% (0.25%) | -1.49% (-0.49%) | 0.5223 |
| Disorganised symptoms | | 2.13% (0.87%) | -0.25% (0.16%) | 0.3093 |
| Years in education | | 10.13% (0.86%) | 7.95% (0.21%) | 1.5773 |
| Educational qualification: GCSE | 11.87% (0.16%) | | | 0.4657 |
| Educational qualification: degree | 4.25% (1.00%) | | | 0.3190 |
| Age at psychosis onset | | 27.76% (0.53%) | 26.00% (0.00%) | 7.5934 |

Note: (a) Tjur's $R^2$ is an alternative to other pseudo-$R^2$ values like Nagelkerke's $R^2$ or Cox-Snell $R^2$ and can be read like any other (pseudo-)$R^2$ value (4). (b) RMSE is the square root of the variance of the residuals and indicates the absolute fit of the model to the data (difference between observed data to model's predicted values). It can be interpreted as the standard deviation of the unexplained variance, and is in the same units as the response variable. Lower values indicate better model fit. (c) (SNP) is the difference between the variance explained by the full model and the variance explained by a covariate-only model and can thus be interpreted as the variance explained by the SNP alone. (d) [1] Adjusted for age at first interview, gender, and principal components (PC)1-5, 6, and 9.

Table 13. Performance metrics for regression models using the observed/imputed data from the Cardiff SibPairs sample (N=148) and rs13107325 allele count as the independent variables.

| | **rs13107325[1]** | | | |
|---|---|---|---|---|
| | **Tjur's $R^2$ (SNP)** | **$R^2$ (SNP)** | **$R^2$ adj. (SNP)** | **RMSE** |
| Three-factor symptom dimensions | | | | |
| Positive symptoms | | 15.93% (3.04%) | 11.09% (2.56%) | 0.2477 |
| Negative symptoms of diminished expressivity | | 11.17% (0.03%) | 6.06% (-0.64%) | 0.5311 |
| Disorganised symptoms | | 17.56% (2.20%) | 12.82% (1.68%) | 0.3588 |
| Years in education | | 17.54% (0.51%) | 12.79% (-0.08%) | 0.8896 |
| Educational qualification: GCSE | 12.22% (0.92%) | | | 0.4632 |
| Educational qualification: degree | 31.65% (1.05%) | | | 0.2346 |
| Age at psychosis onset | | 25.05% (0.39%) | 20.73% (0.15%) | 6.4010 |

Note: (a) Tjur's $R^2$ is an alternative to other pseudo-$R^2$ values like Nagelkerke's $R^2$ or Cox-Snell $R^2$ and can be read like any other (pseudo-)$R^2$ value (4). (b) RMSE is the square root of the variance of the residuals and indicates the absolute fit of the model to the data (difference between observed data to model's predicted values). It can be interpreted as the standard deviation of the unexplained variance, and is in the same units as the response variable. Lower values indicate better model fit. (c) (SNP) is the difference between the variance explained by the full model and the variance explained by a covariate-only model and can thus be interpreted as the variance explained by the SNP alone. (d) [1] Adjusted for age at first interview, gender, and principal components (PC)1-5, except for the degree-dependent variable model where PC3 and PC5 was removed due to multicollinearity.

#### *1. Phenotypes*

##### *1.1. Psychotic experiences*

Table 14. A copy of UK Biobank questions used to define psychotic experiences, taken from the Mental health web-based questionnaire Version 1.3.

| **INTRO7 The next set of questions is about unusual experiences that you may have had, like seeing visions or hearing voices. We believe that these things may be quite common, but we don't know for sure. So please take your time and think carefully before answering:** | | | |
|---|---|---|---|
| F1 | 20471 | Did you ever see something that wasn't really there that other people could not see?<br>Please do not include any times when you were dreaming or half-asleep or under the influence of alcohol or drugs. | [Choose one from]<br>- 01 Yes<br>- 00 No<br>- NA Do not know<br>- DA Prefer not to answer |
| F2 | 20463 | Did you ever hear things that other people said did not exist, like strange voices coming from inside your head talking to you or about you, or voices coming out of the air when there was no one around?<br>Please do not include any times when you were dreaming or half-asleep or under the influence of alcohol or drugs. | [Choose one from]<br>- 01 Yes<br>- 00 No<br>- DA Prefer not to say<br>- NA Don't know |
| F3 | 20474 | Did you ever believe that a strange force was trying to communicate directly with you by sending special signs or signals that you could understand but that no one else could understand (for example through the radio or television)?<br>Please do not include any times when you were dreaming or half-asleep or under the influence of alcohol or drugs. | [Choose one from]<br>- 01 Yes<br>- 00 No<br>- NA Do not know<br>- DA Prefer not to answer |
| F4 | 20468 | Did you ever believe that that there was an unjust plot going on to harm you or to have people follow you, and which your family and friends did not believe existed?<br>Please do not include any times when you were dreaming or half-asleep or under the influence of alcohol or drugs. | [Choose one from]<br>- 01 Yes<br>- 00 No<br>- NA Do not know<br>- DA Prefer not to answer |

##### *1.2. Cognitive ability*

Table 15. The number of participants in the UK Biobank sample, prior to the exclusion of outliers, who completed each cognitive test, *for the first time,* at each of the consecutive assessment visits points.

| **Cognitive Test** | **Cognitive Domain** | **A1 (%)** | **A2 (%)** | **A3 (%)** | **A4 (%)** | **Online (%)** | **Total (%)** | **Direction of test which indicates a higher ability** |
|---|---|---|---|---|---|---|---|---|
| Fluid Intelligence | Reasoning | 116177 (32.72) | 11761 (3.31) | 17968 (5.06) | 213 (0.06) | 44222 (12.46) | 190341 (53.61) | Positive |
| Matrix Pattern Completion | Non-verbal reasoning | NA | 13196 (3.72) | 1143 (0.32) | NA | NA | 14339 (4.04) | Positive |
| Numeric Memory | Working memory | 37732 (10.63) | NA | 22301 (6.28) | 498 (0.14) | 62347 (17.56) | 122878 (34.61) | Positive |
| Pairs Matching | Declarative memory | 354838 (99.94) | 231 (0.07) | 0 | 0 | 0 | 355069 (100) | Negative |
| Reaction time | Processing speed | 353799 (99.64) | 23 (0.01) | 39 (0.01) | 0 | NA | 353861 (99.66) | Negative |
| Symbol Digit Substitution | Processing speed | NA | NA | 24612 (6.93) | 570 (0.16) | 74955 (21.11) | 100137 (28.20) | Positive |
| Tower Rearranging | Executive function | NA | NA | 24405 (6.87) | 579 (0.16) | NA | 24984 (7.04) | Positive |
| Trail Making Test A | Processing speed | NA | NA | 24564 (6.92) | 2926 (0.83) | 77634 (21.87) | 105124 (29.61) | Negative |
| Trail Making Test B | Executive function | NA | NA | 23951 (6.75) | 2890 (0.81) | 77453 (21.81) | 104294 (29.37) | Negative |
| g | Multiple | NA | NA | NA | NA | NA | 53606 (15.10) | Positive |

Note: (a) The cognitive domain is taken from Fawns-Ritchie and Dreary (2020)[1]. (b) Percentages use the total number of UK Biobank participants identified for this study as the denominator (N=355,069) and are rounded to two decimal places. (c) NA is used to indicate when a test was not administered at a study visit, 0 when no participants in the study sample completed the test for the first time at a study visit. (d) Tests in bold are those which contributed to the g score.

Abbreviations: A1, initial assessment visit (2006-2010); A2, first repeat assessment visit (2012-13); A3, imaging visit (2014+); A4, first repeat imaging visit (2019+).

Table 16. Skewness and kurtosis of each cognitive test, before and after transformation, and the total number of participants after the exclusion of outliers in the UK Biobank sample**.**

| **Cognitive test** | **Cognitive domain** | **N** | **Transformation** | **Skewness (before transformation)** | **Kurtosis (before transformation)** |
|---|---|---|---|---|---|
| A1 Fluid Intelligence | Reasoning | 116177 | None | 0.17 | -0.14 |
| A2 Matrix Pattern Completion | Reasoning | 13195 | None | -0.31 | 0.11 |
| Online Numeric Memory | Working memory | 62347 | None | -0.37 | 1.12 |
| A1 Pairs Matching | Declarative memory | 354832 | Log(1 + x) | -0.38 (2.36) | 0.20 (28.51) |
| A1 Reaction Time | Processing speed | 353108 | Log(1 + x) | 0.73 (1.74) | 1.41 (6.87) |
| Online Symbol Digit Substitution | Processing speed | 74950 | None | -0.35 | 0.88 |
| A3 Tower Rearranging | Executive function | 24405 | None | -0.14 | -0.09 |
| Online Trail Making Test A | Processing speed | 77544 | Log(1 + x) | 0.68 (3.40) | 0.78 (67.33) |
| Online Trail Making Test B | Executive function | 77400 | Log(1 + x) | 0.45 (2.23) | 0.28 (15.48) |

Abbreviations: A1, initial assessment visit (2006-2010); A2, first repeat assessment visit (2012-13); A3, imaging visit (2014+); A4, first repeat imaging visit (2019+).

Table 17. Pearson correlation coefficients between the cognitive tests in the UK Biobank sample.

| **Cognitive Test** | | **N (overlap)** | **r** | **(95% CI)** | **p** |
|---|---|---|---|---|---|
| A1 Fluid intelligence | A2 Matrix Pattern Completion | 4266 | 0.32 | (0.29, 0.35) | 7.27e-101*** |
| | Online Numeric Memory | 18739 | 0.30 | (0.29, 0.31) | < .001*** |
| | A1 Pairs Matching | 116177 | -0.21 | (-0.21, -0.20) | < .001*** |
| | A1 Reaction Time | 115620 | -0.15 | (-0.16, -0.15) | < .001*** |
| | Online Symbol Digit Substitution | 28403 | 0.30 | (0.29, 0.31) | < .001*** |
| | A3 Tower Rearranging | 7938 | 0.31 | (0.29, 0.33) | 1.6eE172*** |
| | Online Trail Making Test A | 29028 | -0.22 | (-0.23, -0.20) | 2.44e-301*** |
| | Online Trail Making Test B | 28987 | -0.39 | (-0.39, -0.38) | < .001*** |
| A2 Matrix Pattern Completion | Online Numeric Memory | 36 | 0.21 | (-0.13,0.50) | 0.659 |
| | A1 Pairs Matching | 13157 | -0.18 | (-0.20, -0.16) | 2.79e-95*** |
| | A1 Reaction Time | 13169 | -0.16 | (-0.18, -0.15) | 5.41e-80*** |
| | Online Symbol Digit Substitution | 16 | 0.32 | (-0.21, 0.70) | 0.659 |
| | A3 Tower Rearranging | 13070 | 0.32 | (0.31, 0.34) | 8.86e-313*** |
| | Online Trail Making Test A | 5957 | -0.25 | (-0.27, -0.22) | 4.96e-82*** |
| | Online Trail Making Test B | 5852 | -0.36 | (-0.38, -0.34) | 3.77e-178*** |
| Online Numeric Memory | A1 Pairs Matching | 62245 | -0.1 | (-0.11, -0.10) | 1.20e-147*** |
| | A1 Reaction Time | 62235 | -0.08 | (-0.09, -0.07) | 7.96e-83*** |
| | Online Symbol Digit Substitution | 60671 | 0.23 | (0.22, 0.23) | < .001*** |
| | A3 Tower Rearranging | 77 | 0.23 | (0.00, 0.42) | 0.219 |
| | Online Trail Making Test A | 53763 | -0.20 | (-0.21, -0.19) | < .001*** |
| | Online Trail Making Test B | 53786 | -0.31 | (-0.32, -0.30) | < .001*** |
| A1 Pairs Matching | A1 Reaction Time | 352874 | 0.14 | (0.13, 0.14) | < .001*** |
| | Online Symbol Digit Substitution | 74839 | -0.22 | (-0.23, -0.21) | < .001*** |
| | A3 Tower Rearranging | 24339 | -0.19 | (-0.21, -0.18) | 6.02e-204*** |
| | Online Trail Making Test A | 77392 | 0.15 | (0.14, 0.16) | < .001*** |

| | | | | | |
|---|---|---|---|---|---|
| | Online Trail Making Test B | 77250 | 0.22 | (0.22, 0.23) | < .001*** |
| A1 Reaction Time | Online Symbol Digit Substitution | 74796 | -0.26 | (-0.26, -0.25) | < .001*** |
| | A3 Tower Rearranging | 24358 | -0.16 | (-0.17, -0.15) | 6.78e-135*** |
| | Online Trail Making Test A | 77401 | 0.20 | (0.20, 0.21) | < .001*** |
| | Online Trail Making Test B | 77260 | 0.24 | (0.24, 0.25) | < .001*** |
| Online Symbol Digit Substitution | A3 Tower Rearranging | 30 | -0.02 | (-0.38, 0.34) | 0.91 |
| | Online Trail Making Test A | 64309 | -0.51 | (-0.52, -0.51) | < .001*** |
| | Online Trail Making Test B | 64327 | -0.59 | (-0.59, -0.58) | < .001*** |
| A3 Tower Rearranging | Online Trail Making Test A | 10943 | -0.24 | (-0.25, -0.22) | 1.62e-136*** |
| | Online Trail Making Test B | 10779 | -0.35 | (-0.36, -0.33) | 6.43e-303*** |
| Online Trail Making Test A | Online Trail Making Test B | 77315 | 0.66 | (0.66, 0.66) | < .001*** |

Note: (a) P-value adjustment method: Holm (1979). (b) Observations: N=16-35287. (c) Tests used to make g are in bold. (d) ome p-values were too small to compute in R.
Abbreviations: A1, initial assessment visit (2006-2010); A2, first repeat assessment visit (2012-13); A3, imaging visit (2014+); A4, first repeat imaging visit (2019+).

Table 18. The first four principal components from the principal component analysis of online numeric memory (reverse scored), A1 reaction time, A1 pairs matching, and online trail-making test (TMT) B in the UK Biobank sample.

| | PC1 (g score) | PC2 | PC3 | PC4 |
|---|---|---|---|---|
| Standard deviation | 1.245 | 0.968 | 0.945 | 0.787 |
| Proportion of Variance | 0.388 | 0.234 | 00.223 | 0.155 |
| Cumulative Proportion | 0.388 | 0.622 | 0.845 | 1.000 |

Table 19. Pearson correlation coefficients between g and fluid intelligence and all the individual cognitive tests in the UK Biobank sample.

| Measure of Cognition | Cognitive Test | N (overlap) | r | (95% CI) | P-value |
|---|---|---|---|---|---|
| g | A1 Fluid intelligence | 16151 | 0.41 | (0.40, 0.43) | <0.001 |
| | A2 Matrix Pattern Completion | 20 | 0.35 | (-0.11, 0.69) | 0.127 |
| | Online Numeric Memory | 53600 | 0.61 | (0.60, 0.61) | <0.001 |
| | A1 Pairs Matching | 53600 | 0.53 | (0.52, 0.54) | <0.001 |
| | A1 Reaction Time | 53600 | 0.52 | (0.52, 0.53) | <0.001 |
| | Online Symbol Digit Substitution | 52539 | 0.53 | (0.52, 0.54) | <0.001 |
| | A3 Tower Rearranging | 46 | 0.25 | (-0.05, 0.50) | 0.099 |
| | Online Trail Making Test A | 53543 | 0.53 | (0.52, 0.53) | <0.001 |
| | Online Trail Making Test B | 53600 | 0.79 | (0.79, 0.79) | <0.001 |
| Fluid Intelligence | A2 Matrix Pattern Completion | 4266 | 0.32 | (0.29, 0.35) | 8.07e-102 |
| | Online Numeric Memory | 18739 | 0.30 | (0.29, 0.31) | <0.001 |
| | A1 Pairs Matching | 116177 | 0.21 | (0.20, 0.21) | <0.001 |
| | A1 Reaction Time | 115620 | 0.15 | (0.15, 0.16) | <0.001 |
| | Online Symbol Digit Substitution | 28403 | 0.30 | (0.29, 0.31) | <0.001 |
| | A3 Tower Rearranging | 7938 | 0.31 | (0.29, 0.33) | 1.25e-173 |
| | Online Trail Making Test A | 29028 | 0.22 | (0.20, 0.23) | 1.53e-302 |
| | Online Trail Making Test B | 28987 | 0.39 | (0.38, 0.39) | <0.001 |

Note: (a) Pearson's product-moment correlation. (b) All scores are z-scores where higher values represents better test performance and higher cognitive ability. (c) Tests used to make g are in bold.
Abbreviations: A1, initial assessment visit (2006-2010); A2, first repeat assessment visit (2012-13); A3, imaging visit (2014+); A4, first repeat imaging visit (2019+).

##### *1.3. Educational attainment*

Table 20. A copy of UK Biobank questions used to define educational attainment, taken from the UK Biobank touch-screen questionnaire: final version.

| | | | | | |
|---|---|---|---|---|---|
| D12 | 6138 | Which of the following qualifications do you have?<br>(You can select more than one) | TOGGLE of 8 choices<br>1 : College or University degree<br>2 : A levels/AS levels or equivalent<br>3 : O levels/GCSEs or equivalent<br>4 : CSEs or equivalent<br>5 : NVQ or HND or HNC or equivalent<br>6 : Other professional qualifications eg: nursing, teaching<br>-7 : None of the above<br>-3 : Prefer not to answer | Require ≥1 choices<br>-7 : is exclusive<br>-3 : is exclusive | A levels/AS levels and equivalent includes the Higher School Certificate<br>O levels/GCSEs and equivalent includes the School Certificate. |

Abbreviations: A Level, General Certificate of Education: Advanced Level; O Level, General Certificate of Education: Ordinary Level; CSE, Certificate of Secondary Education; GCSE, General Certificate of Secondary Education; HNC, Higher National Certificate; HND, Higher National Diploma; NVQ, National Vocational Qualification.

#### *2. Missing data*

Table 21. Proportion of non-missing data in the UKBB Sample (N=355069).

| | **UKBB** | |
|---|---|---|
| | **N** | **%** |
| N | 355069 | 100 |
| Age at interview | 355069 | 100 |
| Gender | 355069 | 100 |
| Any psychotic experience | 115422 | 32.51 |
| A distressing psychotic experience | 111936 | 31.53 |
| Multiple psychotic experiences | 112958 | 31.81 |
| Delusions of persecution | 110712 | 31.18 |
| g | 53600 | 15.10 |
| Fluid intelligence | 116177 | 32.72 |
| Years in education | 240436 | 67.72 |
| Educational qualification: GCSEs | 276995 | 78.01 |
| Educational qualification: degree | 333736 | 93.99 |
| Principle components | 355069 | 100 |

#### *3. Descriptive statistics*

Table 22. Descriptive statistics using the observed data from the UKBB sample.

| Phenotypes | rs13107325 schizophrenia-risk allele count | | | | |
|---|---|---|---|---|---|
| | 0 (N=304253) | 1 (N=48907) | 2 (N=1909) | Total (N=355069) | P-Value[1] |
| Age at interview | | | | | |
| Mean (sd) | 56.92 (7.96) | 56.94 (7.94) | 57.19 (7.96) | 56.92 (7.96) | 0.257 |
| N Missing | 0 | 0 | 0 | 0 | |
| Sex | | | | | |
| Male | 140435 (46.16%) | 22893 (46.81%) | 892 (46.73%) | 164220 (46.25%) | 0.025 |
| Female | 163818 (53.84%) | 26014 (53.19%) | 1017 (53.27%) | 190849 (53.75%) | |
| N Missing | 0 | 0 | 0 | 0 | |
| Any psychotic experience | | | | | |
| Yes | 4657 (4.69%) | 707 (4.55%) | 35 (5.91%) | 5399 (4.68%) | 0.270 |
| No | 94636 (95.31%) | 14830 (95.45%) | 557 (94.09%) | 110023 (95.32%) | |
| N Missing | 204960 | 33370 | 1317 | 239647 | |
| A distressing psychotic experience | | | | | |
| Yes | 1658 (1.72%) | 242 (1.61%) | 13 (2.28%) | 1913 (1.71%) | 0.339 |
| No | 94636 (98.28%) | 14830 (98.39%) | 557 (97.72%) | 110023 (98.29%) | |
| N Missing | 207959 | 33835 | 1339 | 243133 | |
| Multiple psychotic experiences | | | | | |
| Yes | 2549 (2.62%) | 369 (2.43%) | 17 (2.96%) | 2935 (2.60%) | 0.320 |
| No | 94636 (97.38%) | 14830 (97.57%) | 557 (97.04%) | 110023 (97.40%) | |
| N Missing | 207068 | 33708 | 1335 | 242111 | |
| Delusions of persecution | | | | | 0.815 |
| Yes | 587 (0.62%) | 98 (0.66%) | 4 (0.71%) | 689 (0.62%) | |

| | | | | | |
|---|---|---|---|---|---|
| No | 94636 (99.38%) | 14830 (99.34%) | 557 (99.29%) | 110023 (99.38%) | |
| N Missing | 209030 | 33979 | 1348 | 244357 | |
| g | | | | | |
| Mean (sd) | 0.01 (1.00) | -0.03 (1.01) | -0.13 (0.97) | 0.00 (1.00) | < 0.001 |
| N Missing | 258259 | 41579 | 1631 | 301469 | |
| Fluid intelligence | | | | | |
| Mean (sd) | 0.01 (1.00) | -0.04 (1.00) | -0.18 (0.94) | 0.00 (1.00) | < 0.001 |
| N Missing | 204552 | 33065 | 1275 | 238892 | |
| Years in education | | | | | |
| Mean (sd) | 16.65 (2.19) | 16.63 (2.18) | 16.57 (2.03) | 16.64 (2.19) | 0.111 |
| N Missing | 98593 | 15439 | 601 | 114633 | |
| Educational qualification | | | | | |
| No GCSE | 92011 (38.81%) | 15263 (39.73%) | 645 (42.57%) | 107919 (38.96%) | < 0.001 |
| GCSE | 145051 (61.19%) | 23155 (60.27%) | 870 (57.43%) | 169076 (61.04%) | |
| N Missing | 67191 | 10489 | 394 | 78074 | |
| Educational qualification | | | | | |
| No Degree | 190752 (66.68%) | 31010 (67.61%) | 1225 (68.09%) | 222987 (66.82%) | < 0.001 |
| Degree | 95320 (33.32%) | 14855 (32.39%) | 574 (31.91%) | 110749 (33.18%) | |
| N Missing | 18181 | 3042 | 110 | 21333 | |

Note: (a) [1] P-values were computed by the arsenal::tableby function which reports an Analysis of variance (ANOVA) (t-test with equal variances) p-value for continuous variables and a Pearson's chi-square p-value for categorical variables.
Abbreviations: sd, standard deviation.

##### *4. Regression analyses*

Table 23. Regression analyses using the observed data from the UKBB sample (N=355069) and rs13107325 allele count as the independent variable.

| | | **Univariable** | | | | | **Multivariable[1]** | | | | | | |
|---|---|---|---|---|---|---|---|---|---|---|---|---|---|
| | **N** | **Beta/OR** | **Lower 95% CI** | **Upper 95% CI** | **Standard Error** | **P-Value** | **Beta/ OR** | **Lower 95% CI** | **Upper 95% CI** | **Standard Error** | **P-Value** | **P-Value adj.** | |
| Any psychotic experience | 115422 | 0.99 | 0.92 | 1.07 | 0.04 | 0.845 | 1.00 | 0.92 | 1.07 | 0.04 | 0.903 | 0.903 | |
| A distressing psychotic experience | 111936 | 0.96 | 0.85 | 1.09 | 0.06 | 0.568 | 0.97 | 0.85 | 1.09 | 0.06 | 0.588 | 0.661 | |
| Multiple psychotic experiences | 112958 | 0.94 | 0.85 | 1.04 | 0.05 | 0.266 | 0.97 | 0.85 | 1.09 | 0.05 | 0.269 | 0.404 | |
| Delusions of persecution | 110712 | 1.07 | 0.87 | 1.30 | 0.10 | 0.523 | 1.05 | 0.86 | 1.28 | 0.10 | 0.611 | 0.786 | |
| g[2] | 53600 | *-0.05* | *-0.07* | *-0.02* | 0.01 | 1.07e-04 | *-0.05* | *-0.07* | *-0.02* | 0.01 | 1.90e-05 | | |
| g score[2,3] | | | | | | | *-0.05* | *-0.07* | *-0.02* | 0.01 | 1.91e-05 | 8.61E-05 | * |
| Fluid intelligence[2] | 53600 | *-0.05* | *-0.07* | *-0.04* | 0.01 | 5.19e-11 | *-0.05* | *-0.07* | *-0.04* | 0.01 | 4.65e-11 | | |
| Fluid intelligence[2,3] | | | | | | | *-0.05* | *-0.07* | *-0.04* | 0.01 | 3.72e-11 | 3.35e-10 | * |
| Years in education | 240436 | *-0.02* | *-0.05* | *0.00* | 0.01 | 0.041 | *-0.03* | *-0.05* | *0.00* | 0.01 | 0.021 | | |
| Years in education[3] | | | | | | | *-0.03* | *-0.05* | *0.00* | 0.01 | 0.019 | 0.035 | * |
| Educational qualification: GCSEs | 276995 | 0.96 | 0.94 | 0.98 | 0.01 | 2.23e-05 | 0.96 | 0.94 | 0.98 | 0.01 | 2.76e-05 | 6.21e-05 | * |
| Educational qualification: degree | 333736 | 0.96 | 0.94 | 0.98 | 0.01 | 4.46e-05 | 0.95 | 0.93 | 0.97 | 0.01 | 2.01e-06 | 6.03e-06 | * |

Note: (a) Ordinary least square regression models. (b) Beta coefficients indicated by italic. (c) [1] Adjusted for age at first interview, biological sex, principal components (PC)1-10, and genotype batch. (d) [2] Additionally adjusted for age at first interview squared. (e) [3] Recalculated standard errors, confidence intervals, and p-values to estimate heteroscedasticity-robust variance. (f) * False Discovery Rate (FDR) adjusted p-value (adjusted for 9 tests) was statistically significant at an alpha level ≤ .05.
Abbreviations: CI, confidence interval; OR, odds ratio.

Table 24. Regression analyses using the observed data from the UKBB sample (N=355069) and rare variant allele count in SLC39A8 as the independent variable.

| | **N** | **N Rare Variants (PTVs/Missense)** | **Beta/OR** | **Lower 95% CI** | **Upper 95% CI** | **Standard Error** | **P-Value** | **P-Value adj.** |
|---|---|---|---|---|---|---|---|---|
| Any psychotic experience | 46651 | 3/1 | 0.00 | NA | 13.29 | 98.34 | 0.930 | 1.395 |
| A distressing psychotic experience | 45272 | 3/1 | 0.00 | NA | $3.38 \times 10^{+03}$ | 160.41 | 0.955 | 1.228 |
| Multiple psychotic experiences | 45651 | 3/1 | 0.00 | NA | $1.85 \times 10^{+10}$ | 255.26 | 0.970 | 1.091 |
| Delusions of persecution | 44749 | 3/1 | 0.00 | NA | $6.34 \times 10^{+34}$ | 693.04 | 0.988 | 0.988 |
| g score[1] | 22476 | 0/2 | *-1.67* | *-2.93* | *-0.42* | 0.64 | 0.009 | 0.081 |
| Fluid intelligence[1] | 54789 | 0/1 | *-2.31* | *-4.25* | *-0.38* | 0.99 | 0.019 | 0.086 |
| Years in education | 89533 | 5/5 | *-0.80* | *-2.16* | *0.55* | 0.69 | 0.244 | 0.549 |
| Educational qualification: GCSEs | 104192 | 6/4 | 0.24 | 0.05 | 0.89 | 0.72 | 0.044 | 0.132 |
| Educational qualification: degree | 126333 | 6/6 | 0.63 | 0.14 | 2.13 | 0.67 | 0.489 | 0.880 |

Note: (a) Ordinary least square regression models. (b) Beta coefficients indicated by italic. (c) Adjusted for the number of synonymous variants each person carries in SLC39A8, age at interview, sex, exome data PC1-10, and sequencing batch. (d) NA indicates that there were not enough data to reliably calculate confidence intervals.
(RV) is the difference between the variance explained by the full model and the variance explained by a covariate-only model and can thus be interpreted as the variance explained by the rare variant allele count alone. (e) [1] Additionally adjusted for age at first interview squared. (f) No False Discovery Rate (FDR) adjusted p-value (adjusted for 9 tests) was statistically significant at an alpha level ≤ .05.
Abbreviations: CI, confidence interval; OR, odds ratio; PTVs, Protein-truncating variants.
