## Supplementary Material Figures for "SLC39A8.p.(Ala391Thr) is associated with poorer cognitive ability: a cross-sectional study of schizophrenia and the general UK population"

### Supplementary Figures

#### Table of Figures

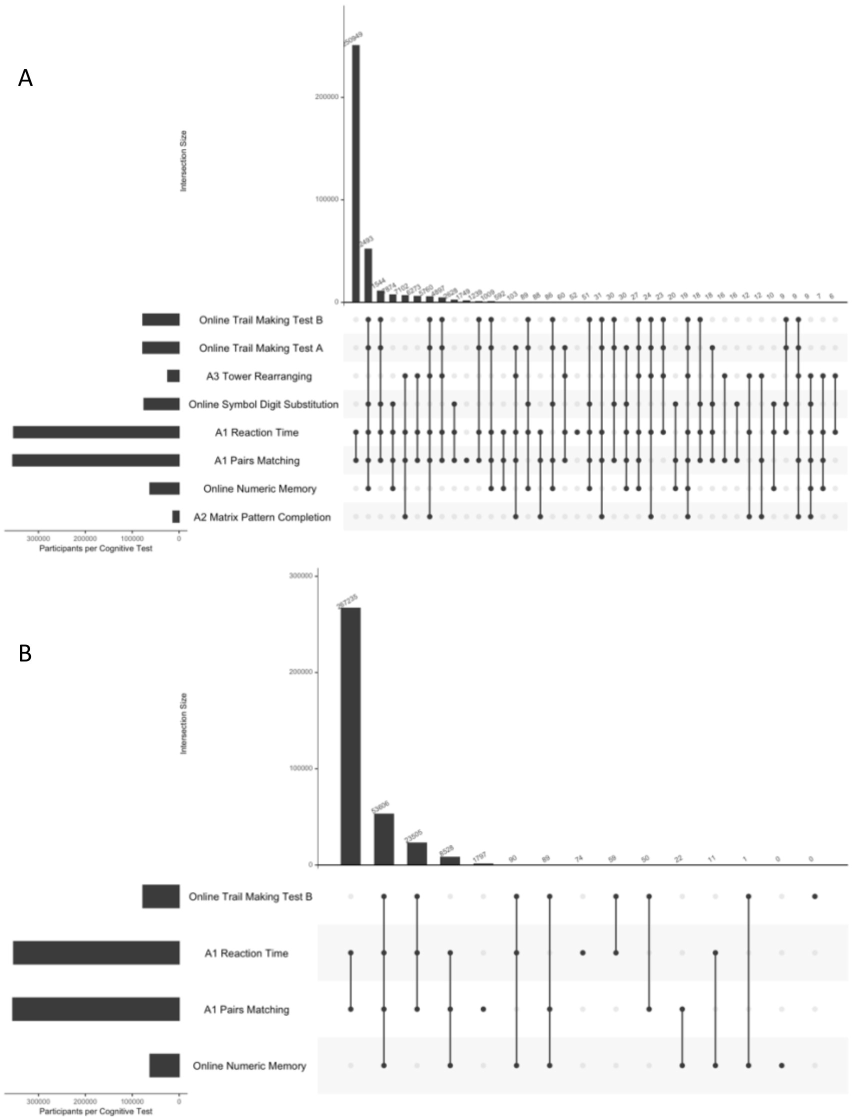

Figure 1. UpSet plot illustrating the number of participants with complete cases data for different combinations of the cognitive tests in UKBB: (A) shows all cognitive test which could have been used to calculate the g score, and (B) shows the four cognitive tests which were used to calculate the g score.

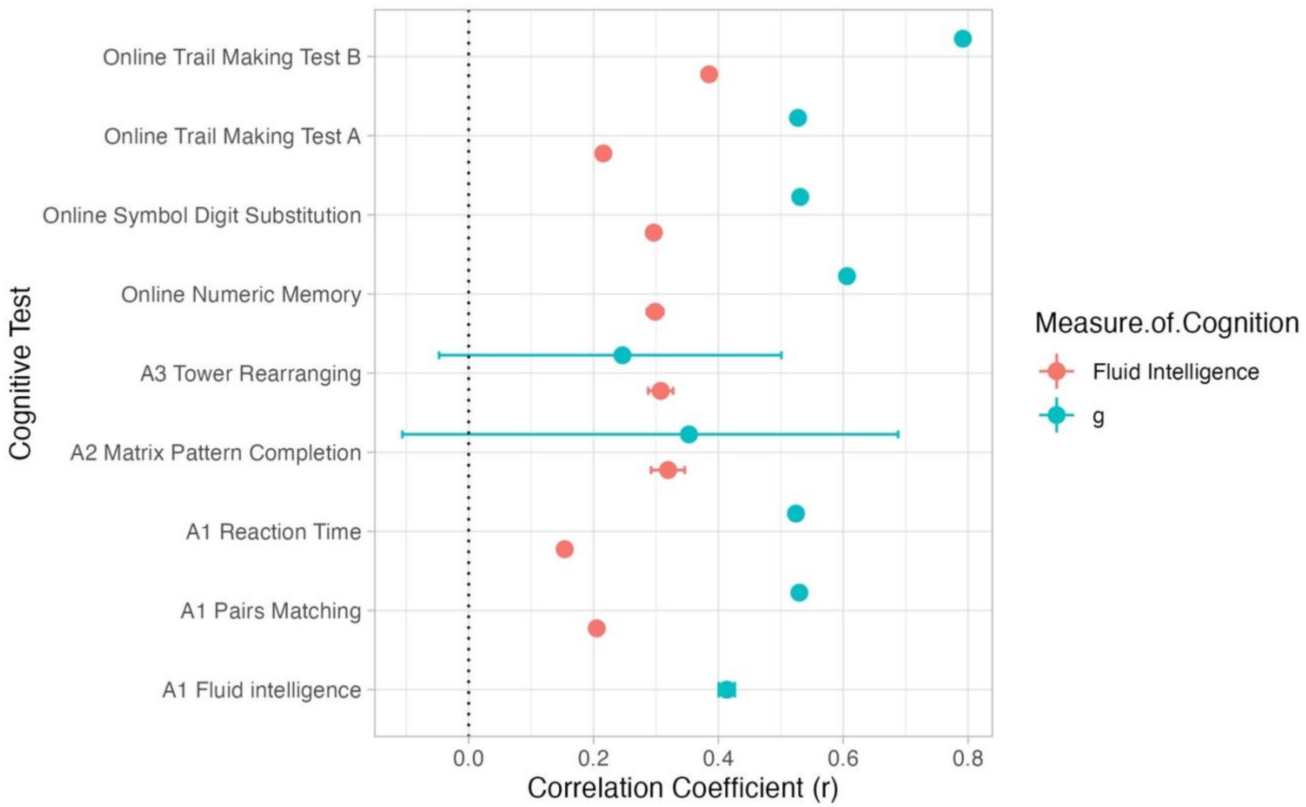

Figure 2. Correlation coefficients between g and fluid intelligence and each of the individual cognitive tests (all measures are z scores where a higher score represents higher cognitive ability) in UKBB. Note, tests used to make g: Online Numeric Memory, A1 Pairs Matching, A1 Reaction Time, and Online Trail Making Test B.

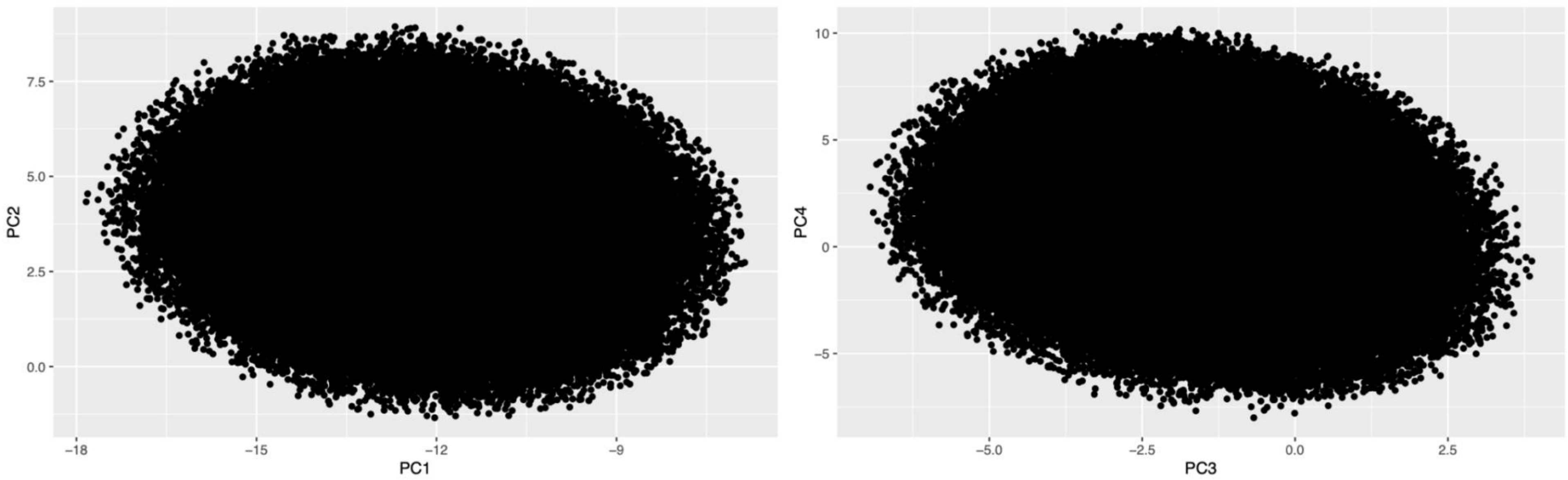

Figure 3. Scatterplots of principal components 1 to 4 for the UKBB subsample after restricting to a genetically homogenous group.
